# Phase 1b dose expansion and translational analyses of olaparib in combination with the oral AKT inhibitor capivasertib in recurrent endometrial, triple negative breast, and ovarian, primary peritoneal, or fallopian tube cancer

**DOI:** 10.1101/2021.04.13.21255421

**Authors:** Shannon N. Westin, Marilyne Labrie, Jennifer K. Litton, Aurora Blucher, Yong Fang, Christopher P. Vellano, Joseph R. Marszalek, Ningping Feng, XiaoYan Ma, Allison Creason, Bryan Fellman, Ying Yuan, Sanghoon Lee, Tae-Beom Kim, Jinsong Liu, Anca Chelariu-Raicu, Tsun Hsuan Chen, Nashwa Kabil, Pamela T. Soliman, Michael Frumovitz, Katheleen M. Schmeler, Amir Jazaeri, Karen H. Lu, Rashmi Murthy, Larissa A. Meyer, Charlotte C. Sun, Anil K. Sood, Robert L. Coleman, Gordon B. Mills

## Abstract

**Background:** Combining poly (ADP-ribose) polymerase (PARP) with phosphatidylinositol-3-kinase (PI3K) pathway inhibitors is supported by strong preclinical rationale. We sought to assess safety and determine a recommended phase 2 dose (RP2D) for PARP inhibitor olaparib combined with the AKT inhibitor, capivasertib, and evaluate molecular markers of response and resistance.

**Methods:** As part of a larger phase 1b trial, we performed a safety lead in of olaparib and capivasertib followed by expansion (n=24) in endometrial, triple negative breast, ovarian, fallopian tube, or peritoneal cancer. Olaparib 300mg orally twice daily and capivasertib orally twice daily on a four day on three day off schedule was evaluated. Two dose levels (DL) were planned: capivasertib 400mg (DL1); capivasertib 320mg (DL-1). Patients underwent biopsies at baseline and after 28 days.

**Findings:** 38 patients were enrolled. 7 (18%) patients had known germline *BRCA1/2* mutations. The first two patients on DL1 experienced dose limiting toxicities (DLTs) of diarrhea and vomiting in absence of maximum supportive care. No DLTs were observed on DL-1 (n=6), therefore, DL1 was re-explored (n=6) with no DLTs, confirming this as RP2D. Most common treatment-related grade 3 or 4 adverse events were anemia (23.7%) and leukopenia (10.5%).

Of 32 subjects evaluable for response, 6 (19%) had partial response (PR) with a PR rate of 44.4% in endometrial cancer. Seven (22%) additional patients had stable disease greater than 4 months. Tumor analysis demonstrated strong correlation between response and immune activity, as well as alterations in cell cycle and DNA damage response genes. Therapy resistance was associated with receptor tyrosine kinase (RTK) and RAS-MAPK pathway activity, as well as metabolism and epigenetics.

**Interpretation:** The combination of olaparib and capivasertib is well tolerated and demonstrates evidence of durable activity in women’s cancers, with particularly promising response in endometrial cancer. Importantly, tumor samples acquired pre and on-therapy can help predict patient benefit.

**Funding:** AstraZeneca, MDACC Moonshots Program, MDACC Support Grant CA016672 NCI SPOREs in Ovarian (CA217685) and Uterine (CA098258) Cancer and a kind gift from the Miriam and Sheldon Medical Research Foundation. AZD5363 was discovered by AstraZeneca subsequent to a collaboration with Astex Therapeutics (and its collaboration with the Institute of Cancer Research and Cancer Research Technology Limited).

## Introduction

Poly (ADP-ribose) polymerase (PARP) inhibitors (PARPi) are now standard of care for breast and ovarian cancer across a number of indications (reviewed in ^1^). Further, these agents may be relevant in endometrial cancers^2^. PARPi demonstrate clear efficacy in tumors harboring aberrations in the homologous recombination (HR) DNA damage repair pathway^1^ with limited activity in tumors that demonstrate innate HR proficiency. Unfortunately, PARPi activity can be of limited duration in the setting of innate resistance or the development of acquired resistance through a diverse array of mechanisms^3^. Thus, there exists a clear unmet clinical need to expand the depth and durability of PARPi responses.

Conversely, activity of single agents targeting the phosphatidylinositol-3-kinase (PI3K) pathway has been limited in women’s cancers. Ovarian cancer demonstrates amplification of *PI3KCA* in 17% of tumors^4^. However, clinical trials of PI3K pathway targeted agents, alone and in combination with chemotherapy, provide no objective benefit in ovarian cancer^5^. In estrogen receptor (ER) positive breast cancer, the PI3K inhibitor alpelisib demonstrated sufficient activity in combination with fulvestrant to receive an indication in *PI3KCA* mutant tumors. In addition, everolimus combined with an aromatase inhibitor improved PFS in patients with hormone-receptor-positive advanced breast cancer^6,7^. Despite having the highest reported rate of aberrations in the PI3K pathway among solid tumors^8^, endometrial cancer demonstrated only modest benefit from PI3K, AKT, or mTORC inhibition as monotherapy^8^.

Combination therapy has arisen as an opportunity to increase the spectrum of patients that benefit from therapy, as well as overcome mechanisms of resistance. Activity of the PI3K pathway has been shown to have a role in tumor cell survival and development of resistance to anti-cancer therapy broadly^9^. We previously found treatment with PARPi led to up-regulation of PI3K pathway members across a number of tumor models^10,11^. Further, inhibition of the PI3K pathway leads to decreased homologous recombination repair through downregulation of *BRCA1/2*, with subsequent increase in DNA damage and sensitivity to PARPi^12^. The AKT serine/threonine protein kinases (AKT1, AKT2, AKT3) are key PI3K pathway downstream mediators and are implicated as oncogenes by constitutive activation through mutation of AKT itself or other constituent of the pathway (loss of PTEN, activating mutation in the catalytic subunit of PI3K (PIK3CA), etc.). Drugs targeting AKT have demonstrated activity in a number of tumor lineages and are being investigated in phase 1/2/3 trials (ie not approved yet).

Xenograft models of HR deficient cancers have demonstrated clear synergy from the combination of PARP inhibition and PI3K inhibition^13^. Objective activity has been validated in early phase clinical trials combining these agents in breast and ovarian cancer^14,15^. However, these trials failed to reach the maximum tolerated olaparib dose due to overlapping toxicity^14,15^. AKT inhibitors (AKTi) have been well-tolerated in clinical studies and demonstrate a different spectrum of toxicity from PARPi suggesting that drug combinations may be tolerated. We sought to evaluate safety and early antitumor activity of the PARPi, olaparib, in combination with two agents targeting the PI3K pathway, capivasertib (AKTi) and vistusertib (mTORC1/2 inhibitor). We report results from the capivasertib arm herein. We explored quality of life and symptom burden in a subset of patients through longitudinal patient reported outcomes. In addition to assessing anti-tumor activity we also collected pre- and on-treatment tumor biopsies to evaluate predictive markers of response to therapy and explore the effect of combination treatment on relevant molecular pathways.

## Methods

### Study Design

This was a phase Ib trial with three non-comparator arms: 1) olaparib + capivasertib given on an intermittent schedule, 2) olaparib and vistusertib given on a continuous schedule, and 3) olaparib and vistusertib given on an intermittent schedule. Patients were enrolled on a given arm based on slot availability. This manuscript describes the olaparib and capivasertib arm, which started with a safety lead in followed by a cohort expansion including ovarian, endometrial or triple negative breast cancer, to further evaluate safety and efficacy and perform translational analyses. The study was conducted at The University of Texas MD Anderson Cancer Center (MDACC) under an IRB-approved protocol. The trial was registered at clinicaltrials.gov as NCT02208375.

### Treatment Plan

Given that a RP2D of the combination of 300 mg olaparib and capivasertib 400 mg, twice a day on a 4/3 schedule had been previously identified in a phase 1 trial across solid tumors^16^ a lead-in phase was planned to obtain additional safety data in women’s cancers. Cycle length was 28 days. Olaparib tablets were administered orally twice daily and capivasertib was administered orally twice daily on an intermittent schedule four days on, three days off. Dose level 1 was olaparib 300mg and capivasertib 400mg. Dose level -1 was olaparib 300mg and capivasertib 320mg. Treatment was continued indefinitely until unacceptable toxicity, disease progression, patient withdrawal, use of prohibited medication, or changes in the patient’s condition that rendered them ineligible for further treatment. The patient population is desbribed in the **Supplemental materials**.

### Patient reported Outcomes

A subset of enrolled patients with endometrial and ovarian cancer participated in a companion study of longitudinal patient reported outcomes (expansion phase) and qualitative interviews (expansion or escalation phase) to explore pattern and severity of symptoms for women who were enrolled in this phase Ib study. The MD Anderson Symptom Inventory (MDASI)-Ovarian cancer module was administered weekly for the first 2 cycles of therapy, then once per cycle, and at study exit^17^. The EQ5D-5L and FACT-ovary were administered at baseline, every 2 cycles, and at study exit^18-20^. See **supplemental table S1** for full schedule of instruments and timepoints and **Supplemental materials** for the description of safety assessment.

### Sample Collection/Molecular Testing

Tissue was obtained through biopsy by interventional radiology at baseline and 28 days after beginning treatment for translational testing. Details of molecular analyses can be found in the **Supplemental materials**.

### Statistical Methods

This study begain as a safety lead in followed by a cohort expansion of the combination of olaparib and capivasertib to further evaluate safety and efficacy and perform translational analyses. Patients who came off study prior to reaching the first evaluation point due to toxicity or disease progression were considered in the DLT analysis. However, patients who came off early for non-medical reasons were not considered in the analysis and were replaced. We used descriptive statistics to summarize the demographic and clinical characteristics of patients, and calculated 90% Bayesian estimation and credible interval for the probability of DLT at each dose level. Adverse events (AE) by CTCAE 4.0 were tabulated by grade, dose level and overall.

Longitudinal symptom burden through the first 8 weeks of treatment was assessed using mixed effect modeling. Covariates included disease site, marital status, education level, body mass index (BMI; obese or non-obese), and age (< or ≥ to 65). Changes from baseline through cycle 2 for PROs other than MDASI were compared using paired t-test and Cohen’s D effect size. Wilcoxon signed-rank test was used when the normality assumption was not met. All statistical procedures were performed using Stata v16 (College Station, TX) or SAS statistical software program for Windows, Version 9.4 (SAS Corporation, Cary, NC). A *p*-value of <0.05 was considered statistically significant.

## Results

### Patient Characteristics

Thirty-eight patients were enrolled on the capivasertib arm of the trial between 9/9/2015 – 2/22/2017, 14 in dose escalation and 24 in dose expansion. Patient characteristics are presented in **Table 1**. Median age was 61 years, patients had a median of 3 (1-7) lines of prior therapy and only one patient had been previously treated with a PARPi. Sixteen patients (42%) had ovarian, 11 (29%) had endometrial, and 11 (29%) had TNBC. Of the ovarian cancer patients, 27% had germline *BRCA* mutations and 87% were platinum-resistant or refractory. Among the endometrial cancer patients, 55% were serous histology and, of those tested, there were no germline *BRCA* mutations. In the TNBC cohort, 27% had a germline *BRCA* mutation.

**Table 1.**
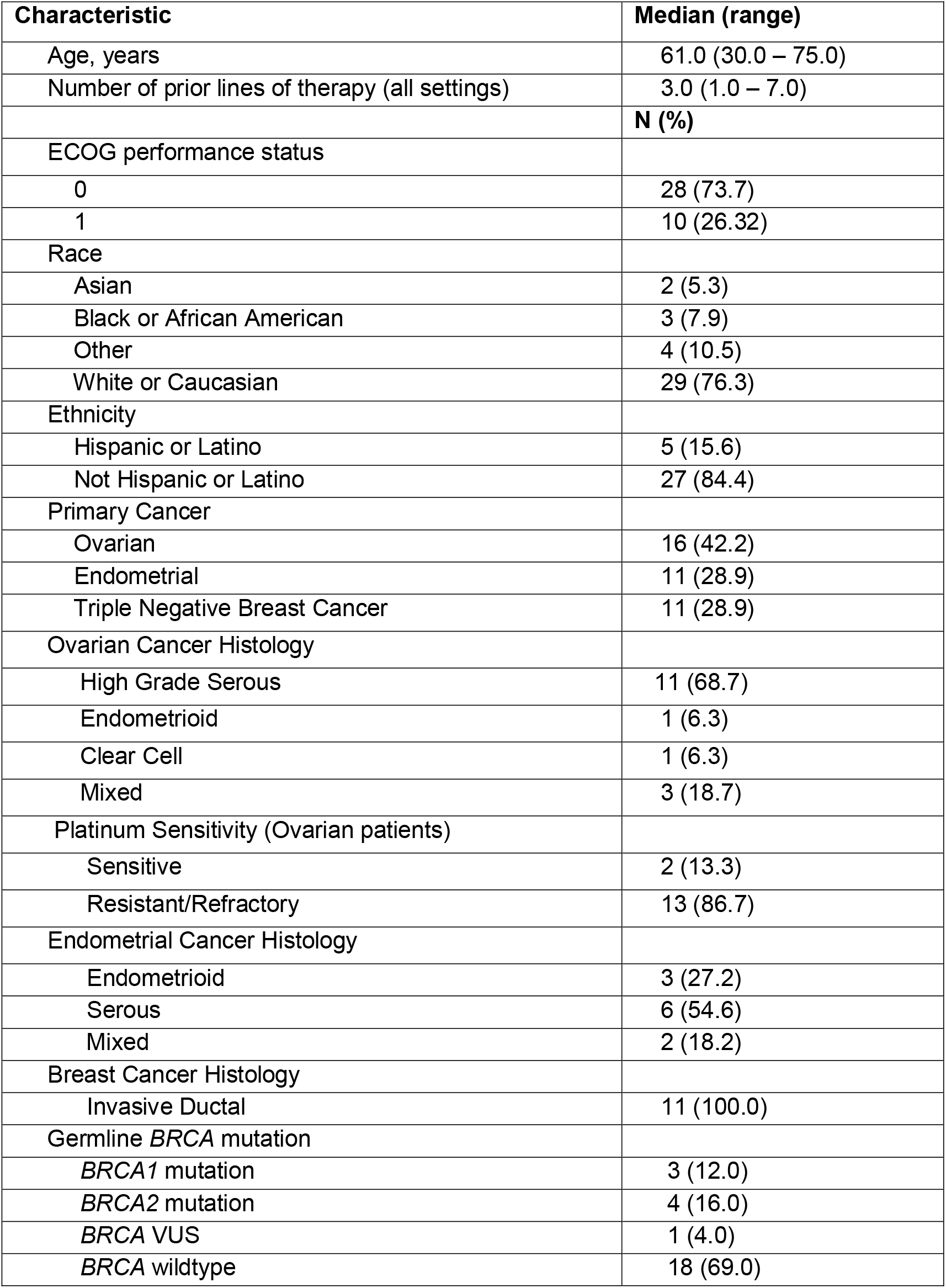
Subject demographics and clinical characteristics (n=38)

### Safety Lead in

**Table 2** demonstrates the dose limiting toxicity estimates for the safety lead in and expansion phases. The first two patients on dose level 1 (olaparib 300mg; capivasertib 400mg) experienced DLTs of diarrhea and vomiting. These toxicities occurred in the absence of maximum supportive care as patients did not report any issue until the adverse events were already grade 3. Thus, 6 patients were treated on dose level -1 (olaparib 300mg; capivasertib 320mg). There were no DLTs on dose level -1, therefore, 6 additional patients were treated on dose level 1 with maximun supportive care. There were no DLTs on the re-explored dose level 1 and this was confirmed as the RP2D. An expansion phase was performed with an additional 24 patients.

**Table 2:**
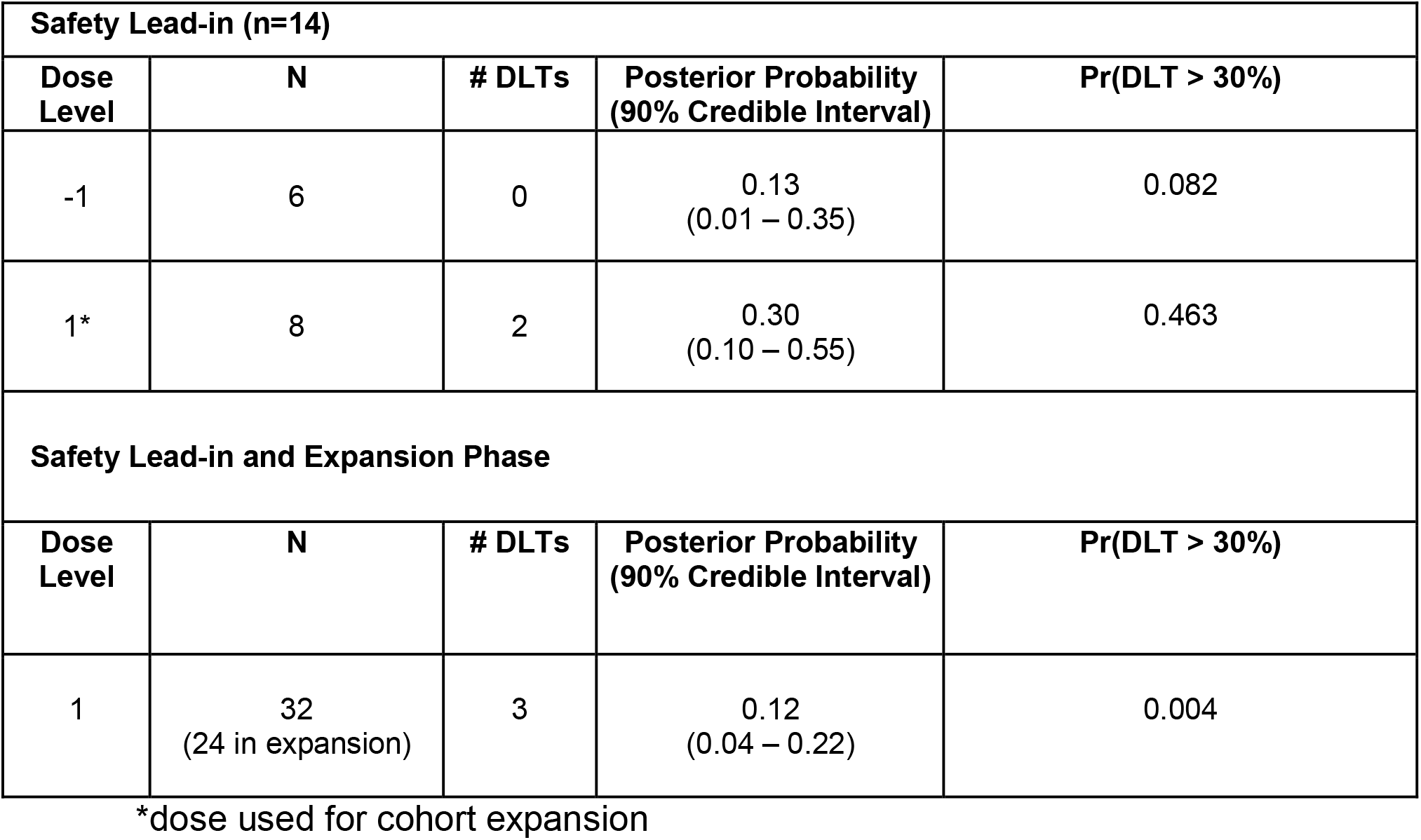
Dose limiting toxicity in safety lead-in and cohort expansion.

### Safety

Thirty-seven (97%) patients experienced at least one treatment-related adverse event **(Table 3)**. The most frequently observed adverse events of any grade were nausea (76%), anemia (63%), diarrhea (61%), elevated creatinine (58%), fatigue (53%), and hyperglycemia (50%). Grade 3/4 adverse events occurred in 19 (50%) patients with some patients demonstrating multiple concurrent adverse events. Through all dose levels, there were 9 (24%) dose interuptions, 8 (21%) dose reductions, and 3 (8%) patients who discontinued due to toxicity.

**Table 3:** All treatment-related adverse events observed in > 10% of subjects*.

| Adverse Event | Grade 1/2 | Grade 3 | Grade 4 | Any Grade |
| --- | --- | --- | --- | --- |
| Nausea | 28 (73.7) | 1 (2.6) | 0 (0.0) | 29 (76.3) |
| Anemia | 15 (39.5) | 9 (23.7) | 0 (0.0) | 23 (63.2) |
| Hyperglycemia | 18 (47.4) | 1 (2.6) | 0 (0.0) | 18 (50.0) |
| Fatigue | 18 (47.4) | 2 (5.3) | 0 (0.0) | 20 (52.7) |
| Elevated creatinine | 21 (55.3) | 1 (2.6) | 0 (0.0) | 22 (57.9) |
| Leukopenia | 10 (26.3) | 4 (10.5) | 0 (0.0) | 14 (36.8) |
| Diarrhea | 21 (55.2) | 2 (5.3) | 0 (0.0) | 24 (60.5) |
| Vomiting | 9 (23.7) | 2 (5.3) | 0 (0.0) | 11 (28.9) |
| Hypertriglyceridemia | 12 (31.6) | 0 (0.0) | 0 (0.0) | 12 (31.6) |
| Mucositis | 11 (28.9) | 0 (0.0) | 0 (0.0) | 11 (28.9) |
| Hypercholesterolemia | 8 (21.1) | 0 (0.0) | 0 (0.0) | 8 (21.1) |
| Thrombocytopenia | 4 (10.5) | 1 (2.6) | 0 (0.0) | 5 (13.1) |
| Headache | 6 (15.8) | 0 (0.0) | 0 (0.0) | 6 (15.8) |
| Neutropenia | 6 (15.8) | 1 (2.6) | 1 (2.6) | 8 (21.1) |
| Constipation | 7 (18.4) | 0 (0.0) | 0 (0.0) | 7 (18.4) |
| Hypokalemia | 9 (23.7) | 0 (0.0) | 0 (0.0) | 9 (23.7) |
| Anorexia | 14 (36.8) | 0 (0.0) | 0 (0.0) | 14 (36.8) |
| Rash | 7 (18.4) | 0 (0.0) | 0 (0.0) | 7 (18.4) |
| Hyponatremia | 6 (15.8) | 1 (2.6) | 0 (0.0) | 7 (18.4) |
| Bladder Infection | 6 (15.8) | 1 (2.6) | 0 (0.0) | 7 (18.4) |
| Abdominal Pain | 5 (13.2) | 0 (0.0) | 0 (0.0) | 5 (13.2) |
| Allergic Reaction | 0 (0.0) | 2 (5.3) | 0 (0.0) | 2 (5.3) |

\* Adverse events less than 10% included if any grade 3/4 toxicity observed. Only causally related adverse events included in this table. 8 patients demonstrated multiple concurrent adverse events.

### Efficacy

Median duration of follow up was 7.4 months (range 0.7 – 37.2). Of 32 subjects evaluable for response, objective response rate was 19% (95% CI: 7.2 – 36.4%). Seven additional patients (22%) had stable disease for greater than 4 months for a clinical benefit rate (CBR) of 41%. **Figure 1** provides additional detail regarding depth and duration of response. Median duration of response was 169 days. Among patients with ovarian cancer, 1 (7%) had partial response and 5 had SD > 4 months (CBR 43%). Of note, 5 of the 6 (83%) ovarian cancer patients with clinical benefit had platinum resistant disease. Objective response rate among endometrial cancer patients was 44% (4/9), with an additional patient achieving SD for > 4 months (CBR 57%). Among TNBC patients, 11% (1/9) achieved objective response and 1 patient (11%) had SD > 4 months (CBR 22%).

**Figure 1.**
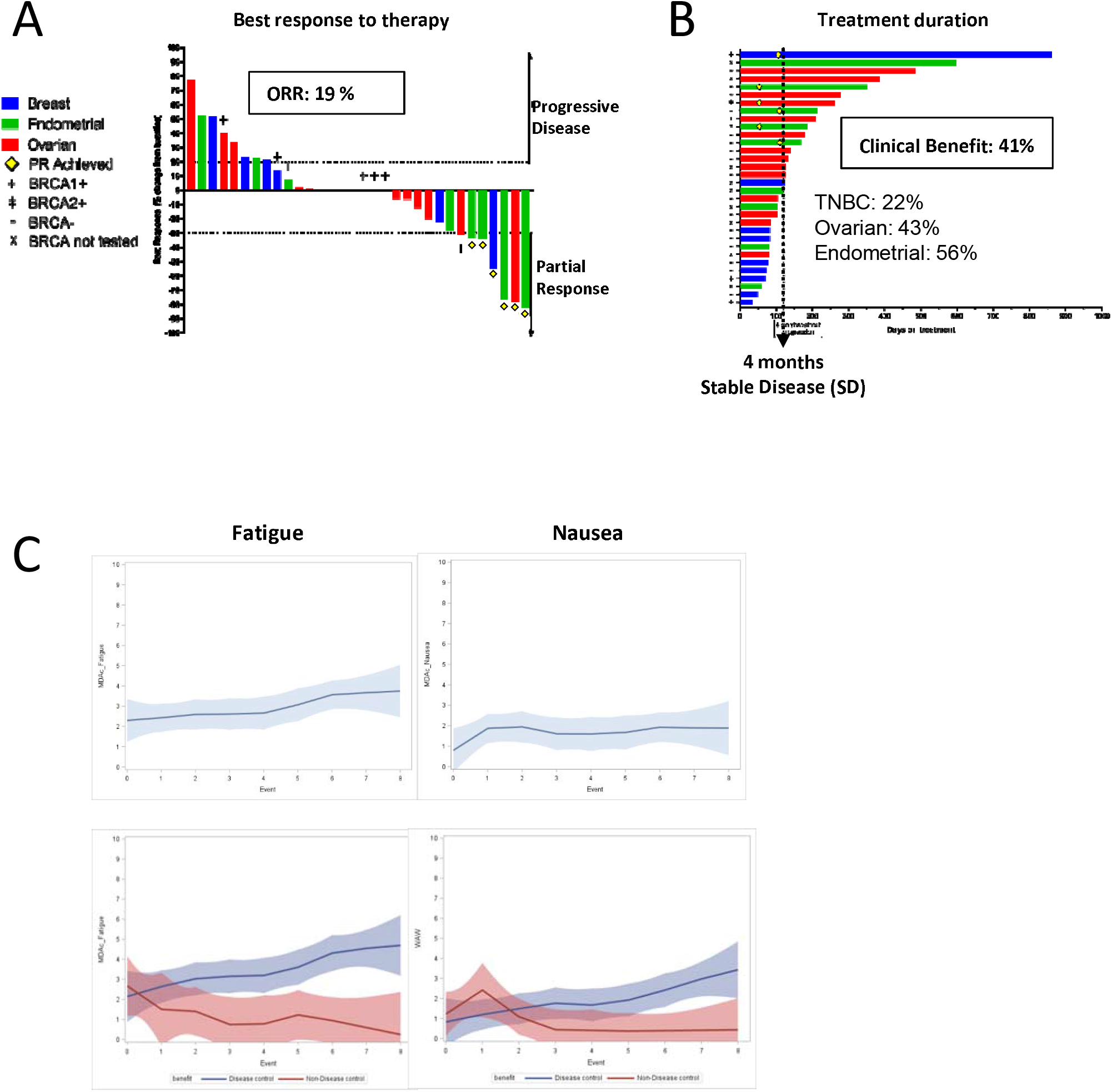
Clinical outcomes. **(A)** Waterfall plot of best response to olaparib and capivasertib among evaluable patients (n=32); **(B)** Swimmer plot of duration on study for all evaluable patients (n=32). **(C)** Longitudinal nausea and fatigue during first 8 weeks of therapy, fatigue between responders and non-responders, interference with physical functioning (walking, activity, work) between responders and non-repsonders.

### Patient Reported Outcomes

15 patients during the expansion phase participated in the companion PRO study, seven women with endometrial cancer and eight with ovarian cancer. Fatigue significantly worsened over time (p=0.04). Those who had clinical benefit reported significantly increased symptom burden of fatigue over time (p<.0001) and increased interference with physical functioning subscore compared to those who did not, p=.004. There was no significant change in nausea from baseline, and overall symptom burden from nausea remained in the mild range **(Figure 1)**.

### DNA analysis

To investigate potential biomarkers of sensitivity and resistance to the drug combination, we combined mutations and small in/dels from WES data (**Figure 2A and supplementary Fig. S1**). Alterations in several genes were found exclusively in the PD group, such as *KRAS* and *FGFR2*. Alterations in the SWI/SNF complex gene *SMARCA2* and cell cycle related genes *CDKN2C* and *CDC27* were found exclusively in the SD and PR groups, as were alterations in DNA repair related gene *ATR. PTEN* mutations were found in two patients, one patient with a PR and one patient with a SD. We assessed association between individual gene alterations and response, and found a total of twelve genes with p-value<0.10 (*KDR, MED12, NTRK2, SPOP, PTEN, FGF5, RARA, ATR, CDC27, PIK3CG, SMARCA2*, and *TSC2*), however, we note that due to small sample size, none of these genes pass multiple correction testing (FDR<0.05). Interestingly, alterations in other PI3K-AKT pathway members were not associated to outcome.

**Figure 2:**
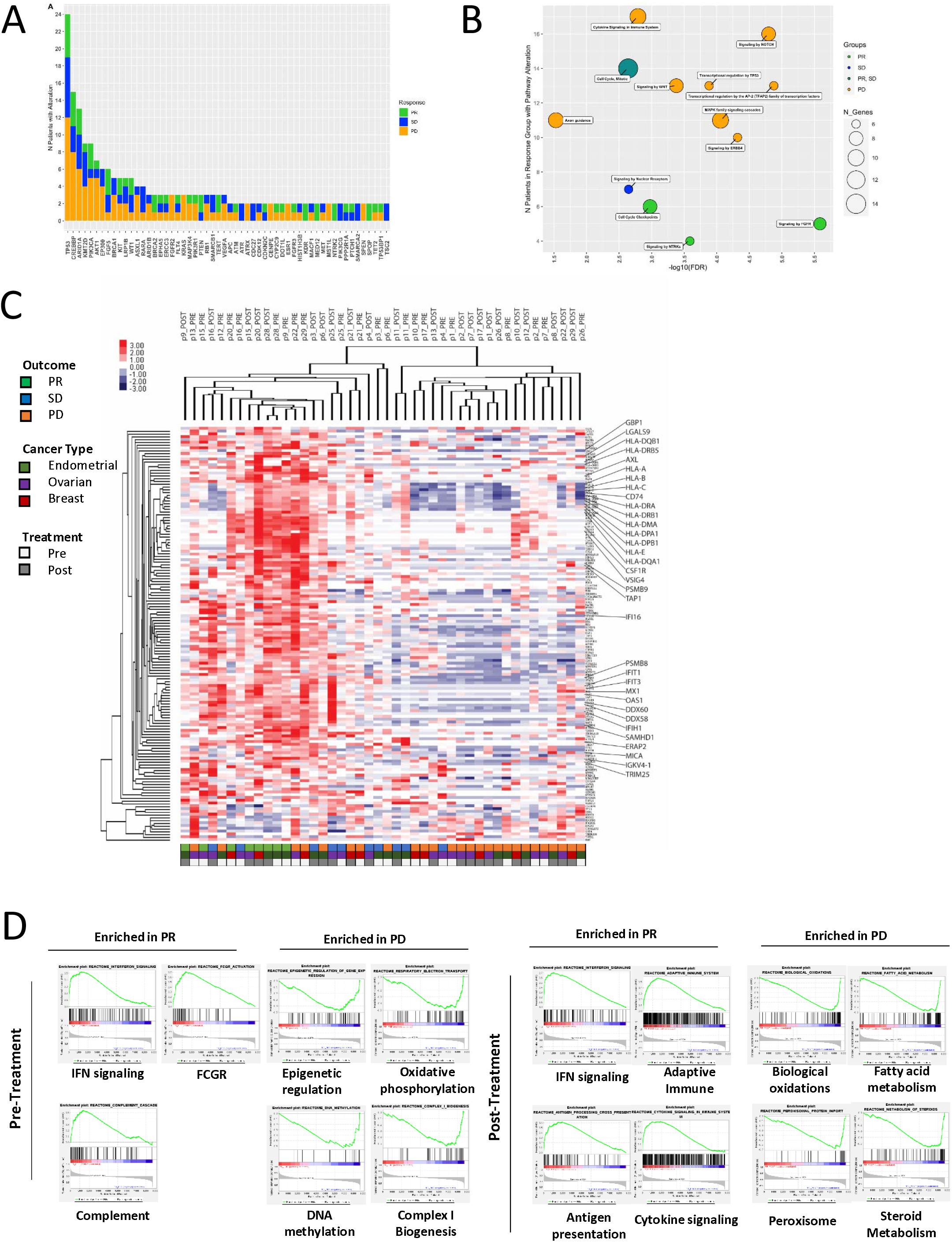
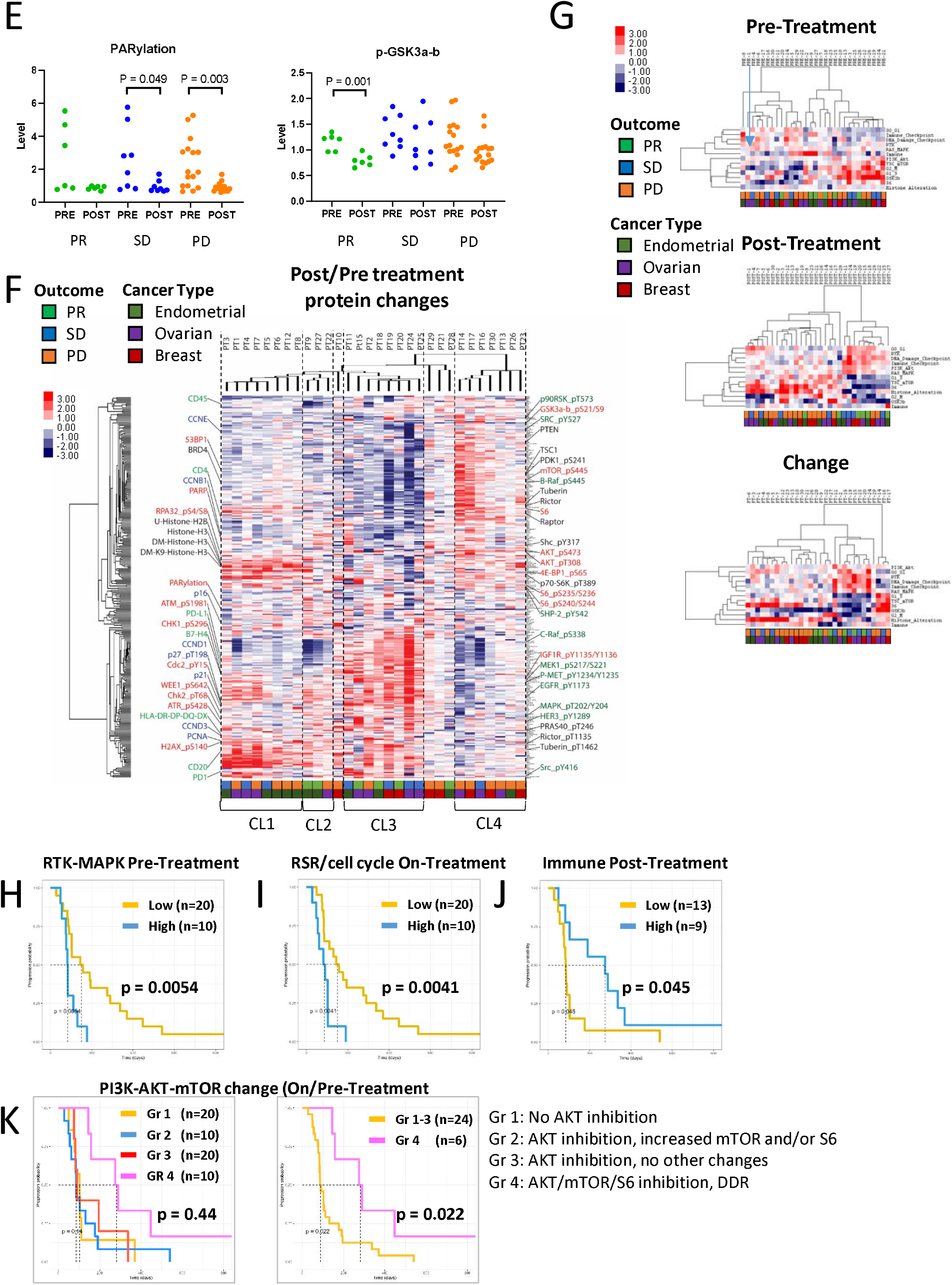
Molecular mechanisms driving the response to therapy. DNA alterations. **(A)** Cohort Distribution for DNA alterations by Response Category. Mutations were aggregated from CLIA-panel and WES, insertion and deletions were called from WES. Alterations were aggregated across PRE and ON-treatment samples for each patient. **(B)** Pathway Enrichment for DNA Alterations, by Response Group. For each response group (PR, SD, and PD), pathways that had an over-representation of altered genes (using both mutation and insertion/deletions) were assessed. Pathway categories are shown on the plot according to number of genes hit in the pathway as well as number of patients with gene alterations, colored according to response group. FDR shown on plot is the minimum of the set of pathways that were collapsed. Full pathway table of top enriched pathways can be found in ***Supplementary Table S2*. Gene expression. (C)** Heatmap representing the hierarchical unsupervised clustering of the RNA expression in pre- and on-treatment sample of all patient. Only the most significant genes between PR and PD patients were included. **(D)** GSEA was performed to compare the expression of genes between PR and PD patients. The most representative pathways enriched in either PR and PD patient’s samples were included. **Protein expression. (E)** RPPA analysis was used to determine protein alteration in the on-treatment samples compared to pre-treatment. Level of total PARylation and phosphorylated GSK3a-b (S21/S9) was measured in the different outcome groups. A paired t-test was used to assess the significant differences between the different groups. P ≤ 0.05 is considered significant. **(F)** Heatmap representing the hierarchical unsupervised clustering of the on to pre-treatment ratio of all proteins and patient samples. Most relevant proteins involved in the cell cycle and DNA damage response, signaling pathways, immune system and epigenetics are identified.**(G)** Pathway analysis comparing pre, on and on to pre change in all samples. The heat maps were constructed using hierarchical unsupervised clustering of both the pathway scores and the samples. A full list of predictors used to calculate the pathways can be found in ***Supplementary Table S3***. Kaplan-Meyer curves were built using the days on treatment for patients with high and low **(H)** RTK and RAS-MAPK pathway activity pre-treatment, **(I)** replication stress and DNA damage response on-therapy and **(J)** immune/INF gene expression on-therapy. Patients with RAS mutation were included in the high RTK-RAS-MAPK group. Only patients with RNA sequencing data were included in the immune/INF Kaplan-Meyer curve. **(K)** Kaplan-Meyer curves showing PI3K-AKT-mTOR inhibition status with and without DNA damage response (DDR). Each patient group was defined based on the pathway scores showed in Figure 2G.

Given the small N observed for both somatic mutations and in/dels, we aggregated gene level alterations to higher functional pathways. Reactome was used to determine whether the presence of SNVs and in/dels of genes in shared pathways was associated with patient response group **(Figure 2B, Supplemental Table S2)**. Aberrant cell cycle checkpoint pathways and signaling of the FGFR or NTRK pathways were highly enriched in the PR group, while “signaling by nuclear receptor pathway” was enriched in the SD group. Strong enrichment of cell cycle and mitosis alterations was found for both PR and SD outcomes. Conversely, several pathways were enriched in the PD group such as the signaling by MAPK family members, NOTCH, WNT, cytokine signaling, and ERBB4 receptor tyrosine kinase (RTK). Transcriptional regulation by TP53 and the AP-2 transcription factor family was also enriched in the PD group.

### RNA Analysis

To further investigate mechanisms of sensitivity and resistance to the drug combination, tumor samples were analyzed by RNA sequencing. A PCA analysis **(Supplemental Figure S2)** did not demonstrate clustering based on tissue sites, suggesting no tissue-dependent response and thus all tumors were analyzed as a set. Furthermore, RNA-based clustering was not dependent on response to therapy or treatment status (pre or on-therapy). The comparison of gene expression between PR and PD tumors showed enrichment of immune related genes in the PR group both pre and on-treatment **(Figure 2C)**. Indeed, most samples from the PD group had markedly lower HLA expression, suggesting reduced antigen presentation by tumor cells. This finding was supported by decreases of other genes involved in immune responses, as shown in **Figure 2C**. GSEA analysis^33^ demonstrated that PR pre-treatment samples were enriched in genes involved in interferon (INF) signaling, complement cascade and FCGR activation. Conversely, genes involved in DNA methylation, epigenetic regulation of gene expression, complex I biogenesis, and respiratory electron transport were enriched in the PD group. In on-treatment samples, immune signatures were strongly associated to PR (interferon signaling, adaptive immune response, antigen presentation, and cytokine signaling) and increased metabolism was associated to PD (biological oxidations, fatty acid and steroid metabolism, peroxisomal protein import) **(Figure 2D)**. To further our findings, a GSVA analysis was used to investigate pathways altered by treatment. As shown in **Supplemental Figure S3**, G2-M DNA damage checkpoint, Myc and E2F targets as well as WNT beta-catenin signaling were decreased on therapy. Because AKTi block cells in G1-S phase, these alterations strongly suggest reduced proliferation, and thus reduced replication stress. Four samples (two PD and two SD) did not have any major alteration of these pathways, suggesting the AKT pathway was not fully inhibited at the doses delivered or at the time of biopsy.

### Protein analysis

Pre- and on-treatment tumors were analyzed by RPPA and a PCA analysis **(Supplemental Figure S2)** revealed clustering based primarily on treatment status. Importantly, no clustering by tissue site was observed. A few samples from the PD group (PT-8, PT-17, PT-21, PT-26) clustered separately from the rest and all of those samples, except for PT-17 also showed a close relationship between pre and on treatment, indicating that the treatment did not induce protein network rewiring that over rode the patient-specific protein network. In Volcano plots **(Supplemental Figure S4)**, PARylation (PAR) was the most downregulated protein, followed by the oxidative stress sensor DJ1, the AKT downstream target p-GSK3a-b (S21/S9) and the replication stress protein RPA32. Proteins upregulated included stress response and apoptotic proteins HSP27, cleaved-caspase-3 and c-IAP2, as well as hormone related p-ERα (S118) and GATA3. Dimethylated lysine 9 Histone-H3 was also increased, supporting activation of the DNA damage response. PARylation and p-GSK3a-b (S21/S9) were used as predictors of target engagement by olaparib and capivasertib. As shown in **Figure 2E**, PARylation was low in all on-treatment samples, although only SD and PD samples had a significant decrease when comparing matched on and pre-treatment samples. This could be explained by a lower PARylation level in the PR samples prior to treatment, although this was not significant. AKT inhibition by capivasertib was more pronounced in the PR groups than in the SD and PD groups, as shown by a significant decreased in p-GSK3a-b (S21/S9).

Unsupervised hierarchical clustering was used to compare expression of proteins in pre- and on-treatment samples as well as changes induced by treatment **(Figure 2F and Supplemental Figure S5)**. Pre-treatment samples showed two major clusters with one enriched in samples from the PD and SD groups (CL2). This cluster was associated with high PI3K-AKT-mTOR activity, as indicated by the strong phosphorylation of AKT, 4E-BP1, GSK3a-b, mTOR and S6. In CL1, a subset of samples from the PD group were enriched in high RTK and RAS-MAPK signaling, as indicated by an elevated phosphorylation of EGFR, MAPK and MEK1. One sample with *KRAS* mutation (p6) and all *FGFR2* mutated (p14, p22, p30) samples were associated with high MAPK pathway activity and PD. Furthermore, two samples from the PR group had strong immune markers (CD4, CD45 and HLAs) consistent with the findings from RNA expression data. The on-treatment samples showed three clusters with two enriched in samples from the PD group (CL2, CL3). CL2 had a high expression of proteins from the mTOR pathway, as well as increased expression and demethylation of histone-H3. CL3 showed high activity of the PI3K-AKT-mTOR pathway with high phosphorylation levels of AKT, GSK3a-b, mTOR and S6. Interestingly, sample 28 from the PR group was part of this cluster. Patient 28 displayed *BRCA2* gene mutation, which could explain sensitivity to treatment in spite of having a high PI3K-AKT-mTORC activity on-treatment. Finally, CL1 was enriched in PR and SD samples and showed a strong RTK and RAS-MAPK activity, which is consistent with a compensatory response to effective AKT inhibition^34,35^. This is markedly different from pre-treatment samples, where high RTK and RAS-MAPK activity was associated with a poor response.

Protein changes induced by treatment resulted in four clusters that separated mostly by outcome. CL1, enriched in PD and SD, showed increased histone alterations (Histone-H3, DM-Histone-H3, Ub-Histone-H2, p-RPA32), consistent with replication stress and DNA damage response. Although PI3K-AKT pathway activity was reduced, these samples had increased mTOR pathway activity (p-4E-BP1 and p-S6) and a moderate RTK-MAPK pathway activation (p-MEK1, p-Met, p-EGFR, p-IGF1R) potentially due to release of feedback inhibition in response to AKT inhibition. These events may be linked as mTORC pathway activity is downstream of RTK-MAPK pathway activity in many epithelial cells^32^. CL2, which contains two PR patients and one PD patient showed inhibition of the PI3K-AKT-mTORC pathway and an induction of DNA damage response as shown by increased p-H2AX and G2-M DNA damage checkpoint proteins p-ATR, p-WEE1 and p-CDC2. CL3 was enriched for both PR and SD patients. This cluster showed the overall strongest PI3K-AKT-mTOR pathway inhibition, as well as increased RAS-MAPK pathway activity potentially due to strong PI3K-AKT pathway inhibition. This cluster also demonstrated activation of several proteins involved in the DNA damage response. Finally, PD enriched CL4 showed increased PI3K-AKT-mTOR pathway activity (p-AKT, p-GSK3a-b, p-mTOR, p-4EBP1 and p-S6) consistent with bypass of the AKT inhibitor.

To investigate the roles of cancer-associated pathways, we used calculated pathway scores from RPPA data **(Figure 2G)**. Unsupervised clustering of these scores demonstrated that although pre-treatment samples did not cluster by outcome, high RTK or RAS-MAPK activity prior to treatment is, in most cases, associated with a poor outcome. In on-treatment samples, the clustering showed that histone alterations and mTOR pathway activity are associated with the PD group. Conversely, in on-therapy biopsies responders showed low AKT activity (p-GSK3b and mTORC) and a high DDR and G0-G1 cell cycle arrest, suggesting a mechanistic response to the drugs. These samples also displayed high RTK, RAS-MAPK, PI3K-AKT (AKT and upstream) signaling activity, suggesting activation of compensatory mechanisms due to inhibition of negative feedback loops. Interestingly, the samples also displayed a high immune checkpoint activity, which again support the involvement of the immune system in the response. Finally, by comparing on to pre-treatment score changes, four types of responses to therapy were observed: 1) inhibition of AKT signaling with an increased mTOR/S6 pathway and histone alteration was associated to bad outcome; 2) inhibition of AKT signaling with almost no change in other pathways, suggesting an indifference to therapy was associated with a poor outcome; 3) increased AKT and mTOR signaling as well as increased G1-S cell cycle phase was associated with therapy resistance; and 4) inhibition of AKT and mTORC/S6 activity as well as a reduced cell cycle progression was associated to good outcome. In responders, DNA damage and immune checkpoints as well as compensatory mechanisms (RTK and MAPK) were activated.

### Biomarkers of response to therapy

Kaplan-Meyer curves were constructed with the different categories of potential biomarkers **(Figure 2H-K, Supplemental Figure S6)**. Integration of the DNA and RNA analysis revealed an association between high RTK and RAS-MAPK pathway activity in pre-treatment samples and a bad outcome (median 83.5 compared to 151.5 days in control patients (p=0.0054)). Patients with high replication stress and DNA damage response in on-treatment samples had reduced time on therapy (median 86 compared to 152 days in control patients (p=0.0041)). Strikingly, patients with a high interferon-pathway and immune signature in on-treatment sample were treated for a longer period of time (275 days) compared to control patients (85 days) (p=0.038). Combined AKT and S6 pathway activation with concurrent induction of the DNA damage was associated with a good outcome (Figure 2K). Taken together, these analysis reveal a potential set of biomarkers that could improve patients selection for this particular combination and help predict outcome.

### PDX model

To investigate independent contributions of olaparib and capivasertib in patient response, we assessed the effects of mono and combination therapy in a PARP-resistant TNBC PDX model **(Figure 3 and Supplemental Figure S6)**. NGS immune deficient mice were treated for five cycles with either vehicle, olaparib, capivasertib, or their combination. Tumor growth monitoring confirmed resistance to olaparib. Capivasertib alone slightly decreased tumor growth, while the combination had a synergistic activity (p<0.001) **(Figure 3A)**. Unsupervised clustering of the protein expression data after five cycles of treatment demonstrated highly conserved protein changes across tumors based on treatment **(Figure 3B)**. The PARP and AKT inhibitors both displayed target engagement, as indicated by decreased PARylation and p-GSK3a-b. Although the olaparib monotherapy group was similar to the vehicle treated group, a slight decrease in G2-M phase proteins (CCNB1 and CDK1) was observed, suggesting a decrease in cell cycle progression. Capivasertib monotherapy triggered several changes with a subset being conserved in the combination therapy group, such as decreased phosphorylation levels of GSK3a-b, S6, CCNB1 and RPA32 and increased expression of p16INK4a, CCND1, p-AKT and p-WEE1. Conversely, some changes induced by capivasertib were reversed by the addition of olaparib. For example, several structural proteins (p-cadherin, d-a-tubulin, b-actin and fibronectin) were decreased following capivasertib monotherapy but not following combination therapy. Others such as PTEN, PDGFR-β and PARylation were increased exclusively in the capivasertib group. The addition of olaparib reversed this phenotype returning levels to that of vehicle treated tumors.

**Figure 3.**
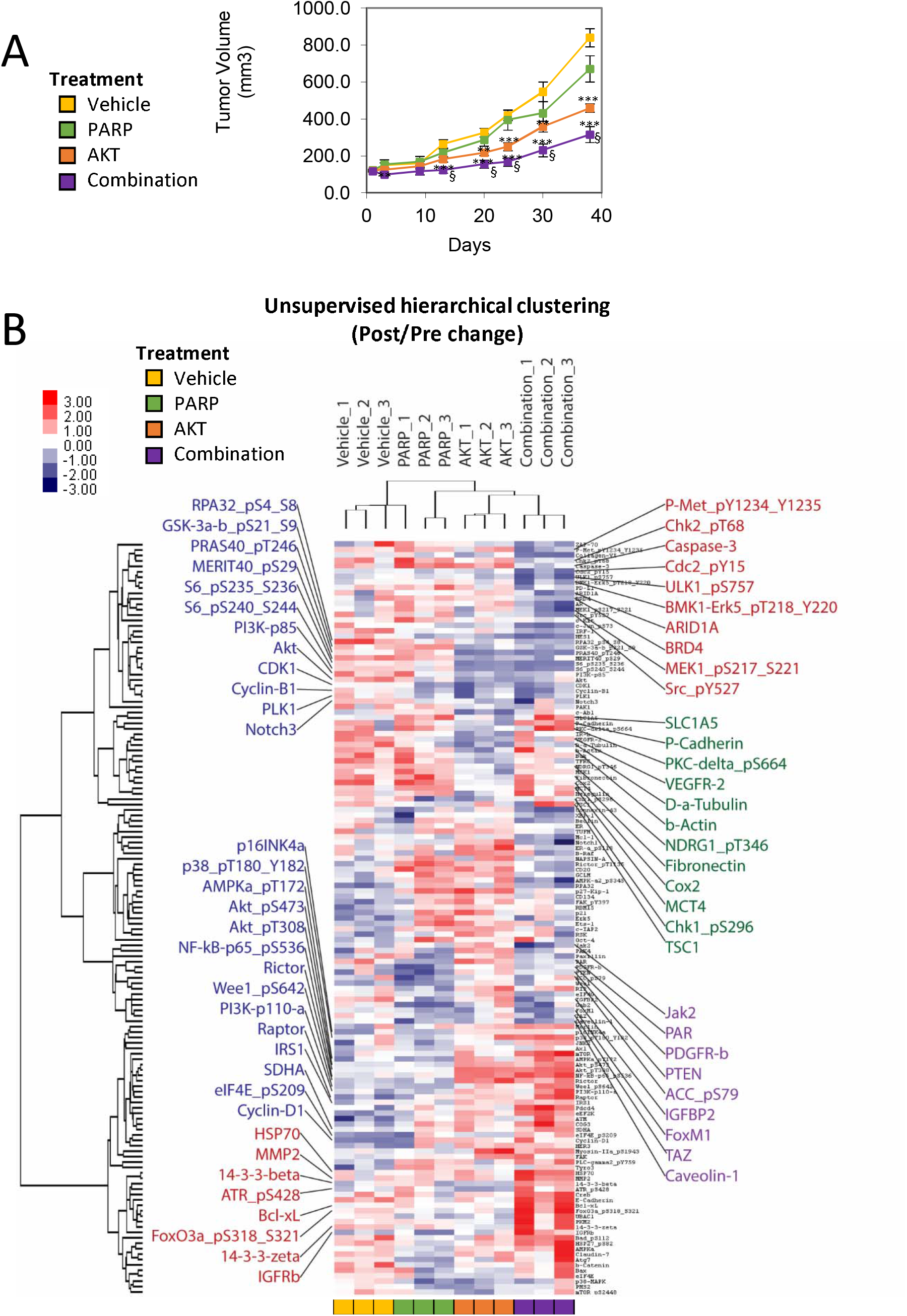
PDX model. A TNBC PDX model was implanted in NGS mice and treated with a vehicle, olaparib, capivasertib or their combination. **(A)** tumor growth was assessed and **(B)** protein were analyzed by RPPA. The heat map representing the hierarchical unsupervised clustering of all proteins with different level of expression across treatment groups. Groups of proteins that were altered by AKT only (green), AKT and the combination (blue), PARP and the combination (purple) and the combination alone (red) were identified. A student t.test was used to compare the tumor growth between control mice and mice treated with olaparib, capivasertib or their combination. **:p<0.01, ***p<0.001. § indicates a significant difference between the tumor size of capivasertib treated mice and mice treated with the combination.

Interestingly, the drug combination altered some proteins independently from olaparib or capivasertib monotherapy. Amongst these changes, we observed an increase in several stress related proteins (HSP70, p-ATR and BCL-xL) and a decrease in the epigenetic mediators BRD4, ARID1A, the DNA damage checkpoint proteins p-chk2, p-cdc2 and signaling molecules p-MET, p-srcpErk5 and p-MEK1. Pathway score analysis revealed that PARP itself did not alter major pathway activity, except for a slight G2-M phase decrease. AKT inhibitor showed a strong decrease of PI3K-AKT-mTOR signaling. The PARP and AKT inhibitor combination blocked both PARylation and PI3K-AKT-mTOR pathway activity. In addition, G2-M phase, total and dimethylated histone-H3, and p-S6 expression were reduced, while the G0-G1 phase increased in combination treated tumors.

## Discussion

This phase 1b study with planned expansions in recurrent ovarian, endometrial and TNBC confirmed the recommended phase 2 dose for olaparib and capivasertib. In future trials, olaparib should be given at a dose of 300mg BID continuously and capivasertib at a dose of 400mg BID on a four day on, three day off schedule. Importantly, there were no unexpected safety signals in this cohort of women’s cancers. Further, there was encouraging clinical activity in a highly pre-treated cohort of patients, including impressive durable responses in women with recurrent endometrial cancer, which is typically resistant to therapy. The extensive translational analyses including analysis of change in protein expression and pathway activity on therapy provide critical molecular insights regarding predictors of response and resistance that could be applied in future trials.

Although adverse events were common on this trial, the majority were grade 1 or 2 and could be mitigated with protocol-directed supportive care. Adverse events were as expected, including known class effects from PARP inhibition and AKT inhibition such as fatigue, gastrointestinal and hematologic toxicity. Our experience with the first two patients on study highlights the need for early intervention for gastrointestinal effects including nausea and diarrhea. After these two early DLTs occurred, our team increased pre-treatment counseling and encouraged patients to report side effects early so that severe toxicity could be avoided.

Overall, the combination of olaparib and capivasertib appeared to be well tolerated from a PRO perspective. Despite frequency of side effects noted such as nausea, anemia and fatigue, the mean symptom burden was in the mild range. Interestingly, patients who received clinical benefit from the combination therapy appeared to have more fatigue and greater interference with physical functioning compared to those who did not receive clinical benefit and experienced disease progression. Given the small numbers, this observation is hypothesis generating and suggestive of more effective target inhibition but warrants further investigation.

Antitumor activity was seen across all tumor types, regardless of presence of *BRCA* mutation or aberrations in the PI3K/AKT pathway. The overall objective response rate and clinical benefit rate are impressive given the high proportion of platinum resistance (87% of ovarian cancer patients) and the low rates of *BRCA* mutations across the study population (0% endometrial cancer, 27% TNBC and 27% ovarian cancer patients). Previous studies of PARP and PI3K inhibition have demonstrated encouraging activity in similar populations, however, prior studies included higher proportions of patients with germline and somatic *BRCA* mutations and did not include endometrial cancers, making it difficult to perform direct comparisons^14,15^.

Previously, activity of single agent PARP inhibition has been most impressive and durable in *BRCA* aberrant disease. Thus, it is important to demonstrate the activity of PARP combinations in patients with *BRCA* wildtype tumors. Indeed, the combination of PARP and AKT had unexpected activity in BRCA wildtype tumors. Further, as more patients are treated with PARPi in the upfront setting in breast and ovarian cancer, there is emergent need to overcome PARP resistance through combination therapy. We have previously shown that the PI3K/AKT pathway is up-regulated in response to PARP inhibition, even after only a short window of treatment^10^. The combination of PARP and AKT inhibition holds promise to reverse or prevent the emergence of PARP resistance, and further studies are necessary to explore this potential. Importantly, this is the first study to show significant activity of PARP inhibition, albeit as part of a combination therapy, in endometrial cancer. It is particularly intriguing that there was clinical activity in this study regardless of endometrial cancer histology or presence of PI3K/AKT pathway aberrations. Moving forward, it will be essential to determine the relative contribution of the PARPi versus the AKT inhibitor in this population. Of note, in prior studies, activity of capivasertib monotherapy was limited to tumors with known PI3K pathway aberrations^36,37^. The TNBC PDX model that we explored in this study helps elicit the contribution of each drug and the synergistic effect of the combination. Indeed, this PARP-resistant model had almost no protein changes with PARPi monotherapy, while AKT inhibition increased DNA damage checkpoint proteins. Importantly, the combination of both drugs drastically reduced tumor growth likely through the induction of major stress responses and decreased cell cycle progression that was not apparent with either agent alone.

By collecting samples pre- and on-therapy from each patient and performing extensive DNA, RNA and protein analysis, we demonstrated that longitudinal analyses can help identify patients likely to benefit from the olaparib and capivasertib combination, as well as identify potentially targetable mechanisms of resistance. We demonstrated, through a pathway-oriented analysis, that patients with cell cycle and DNA damage repair pathway alterations, as well as patients with high immune/INF activity are more likely to respond to therapy. Conversely, patients with *KRAS* mutations or with elevated RTK and RAS-MAPK pathway activity in pretherapy biopsies were resistant to the drug combination. These patients will likely require a different therapy approach. We previously demonstrated that RAS-mutant tumors are sensitive to the combination of PARP and MEK inhibitors, which suggest these patients could benefit from such therapy^11^. There was also a strong association between changes in epigenetic mediators and metabolism with therapy resistance, which suggest a proficient DNA repair mechanism and a possible bypass of the AKT pathway inhibition through mTOR activation. Indeed, previous studies demonstrated that epigenetic events are involved in the resistance to PARPi through the protection of the replication fork and reduced replication stress^38-40^. Further, resistance to AKTi has been previously associated with increased mTOR signaling, mitochondrial biogenesis and oxidative phosphorylation^41-43^. When comparing on-to pre-treatment samples, we observed major protein network rewiring in responders, consistent with the inhibition of negative feedback loops (increased RTK and RAS-MAPK activity)^44^ as well as the induction of a stress response (reduced cell cycle progression and induced DNA damage response). Conversely, several progressing tumors showed signs of incomplete AKT pathway inhibition, potentially as the result of activation of bypass mechanisms. Interestingly, protein data indicate that inhibition of the mTOR axis along with AKTi could improve responses and that the mTOR axis might be a significant contributor to DNA damage repair in response to PARPi^45,46^.

In summary, this study found that the combination of olaparib with capivasertib, demonstrated encouraging activity with acceptable toxicity in women’s cancers. The striking response rates, particularly in endometrial cancer that often is resistant to therapy, support further exploration of the combination. Further, we have identified a series of biomarkers associated with response (cell cycle gene aberration, high IFN signaling, intact antigen presentation, inhibition of AKT pathway activity in on therapy biopsies) and resistance (*KRAS* and *FGFR2* mutations, RTK and RAS-MAPK pathway activity, high epigenetic regulation and oxidative phosphorylation) that, following confirmation in additional studies, could be used to identify and select patients most likely to benefit. Based on our data, treatment genomic evaluation may inform therapeutic decision-making by enabling strategic therapeutic pivoting to agents targeting adaptive responses. Further, although we demonstrated that serial on-therapy biopsies provided the most predictive information for patient outcomes, predictive biomarkers in the pre-treatment sample had considerable ability to identify patients likely to benefit.

## Data Availability

All data referred to in the manuscript will be made available upon reasonable request.

## Supplemental Figures

**Supplemental Figure S1. DNA Alteration. (A)** Cohort Distribution for DNA alterations by Response Category. Mutations were aggregated from CLIA-panel and WES, insertion and deletions were called from WES. Alterations are aggregated across PRE and ON-treatment samples for each patient. Colors indicate response (PR=green, SD=blue, PD= orange). **(B)** Cohort Distribution for DNA alterations by alteration type. Mutations were aggregated from CLIA-panel and WES, insertion and deletions were called from WES. Alterations are aggregated across PRE and ON-treatment samples for each patient. Colors indicate alteration type (deletion= blue, deletion and mutation=dark blue, insertion=light green, insertion and mutation=dark green, mutation= red) **(C)** Cohort Distribution for DNA alterations for selected genes in PI3K/AKT and RAS pathways. Mutations were aggregated from CLIA-panel and WES, insertion and deletions were called from WES. Alterations are aggregated across PRE and ON-treatment samples for each patient. Colors indicate response (PR=green, SD=blue, PD= orange). **(D)** Cohort Distribution for DNA Mutations for Selected Genes in PI3K/AKT and RAS pathways, shown according to Functional Annotation. Mutations were aggregated from CLIA-panel and WES, functional annotation calls as described in Methods. Mutations are aggregated across PRE and ON-treatment samples for each patient. Colors indicate functional annotation (pathogenic=light red, variant of unknown significance (VUS)= grey).

**Supplemental Figure S2. Principal component analysis**. PCA analysis showing the distribution of all samples RNA (left panel) and protein (right panel) expression by treatment status, outcome and tissue of origin (site).

**Supplemental Figure S3. GSVA analysis of the RNAseq data**.

**Supplemental Figure S4. Volcano plot**. Volcano plot showing the most downregulated and upregulated proteins in the on-treatment sample compared to pre-treatment. Proteins with a p-value ≤ 0.05 are in blue and proteins with a p-value ≤ 0.01 and at least 10% fold-change were identified.

**Supplemental Figure S5. RPPA analysis**. Heatmap representing the hierarchical unsupervised clustering of **(A)** pre- and **(B)** on-treatment protein expression data from all patient samples. Proteins involved in the cell cycle and DNA damage response, signaling pathways, immune system and epigenetics are identified.

**Supplemental Figure S6. Kaplan-Meyer curves of the days on treatment**. Group of patients were compared by treatment status (pre, on-therapy) and changes induced by therapy for the PI3K pathway activity, the immune score and the RTK-MAPK pathway activity.

**Supplemental Figure S7. Adaptive response to olaparib, capivasertib and their combination in a TNBC PDX model**. Protein pathway activity in a TNBC PDX model treated with vehicle (yellow), olaparib (green), capivasertib (orange) and their combination (purple) for 5 cycles. The heat map represent the unsupervised clustering of the pathway scores.

