## Supplemental Figures for "Phase 1b dose expansion and translational analyses of olaparib in combination with the oral AKT inhibitor capivasertib in recurrent endometrial, triple negative breast, and ovarian, primary peritoneal, or fallopian tube cancer"

### Slide 1
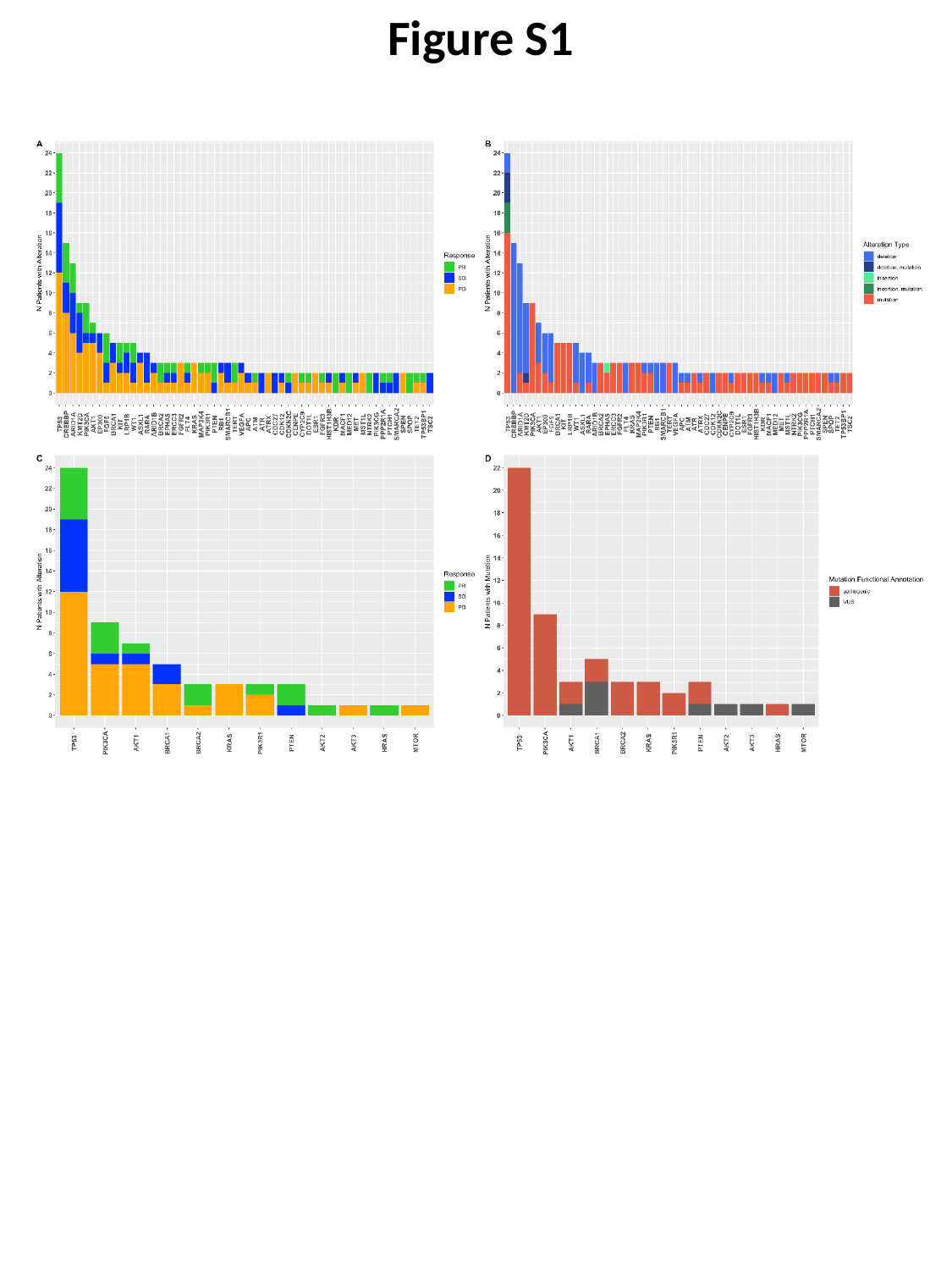

Figure S1

### Slide 2
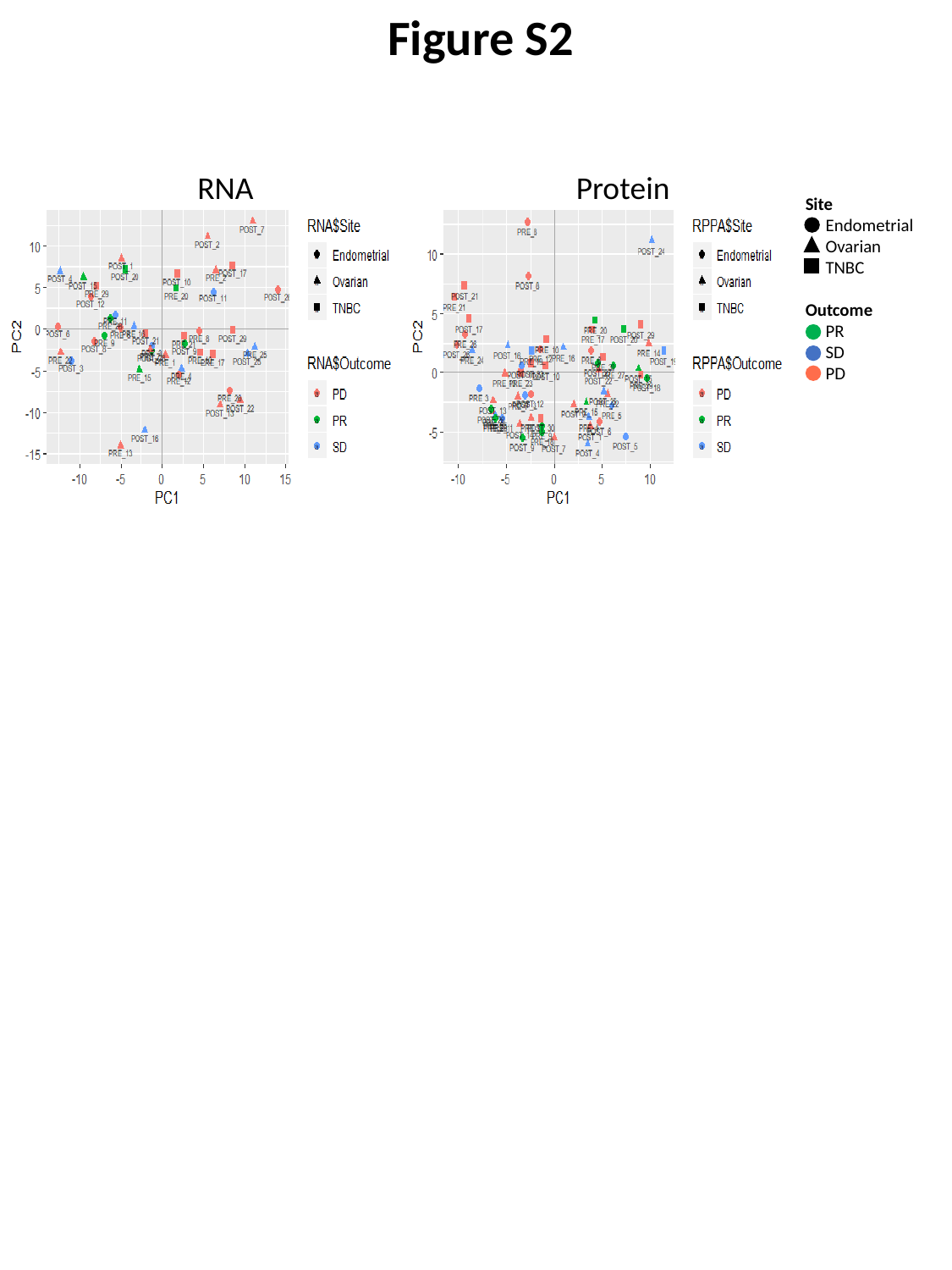

Figure S2
RNA
Protein
Site
 Endometrial
 Ovarian
 TNBC
Outcome
 PR
 SD
 PD

### Slide 3
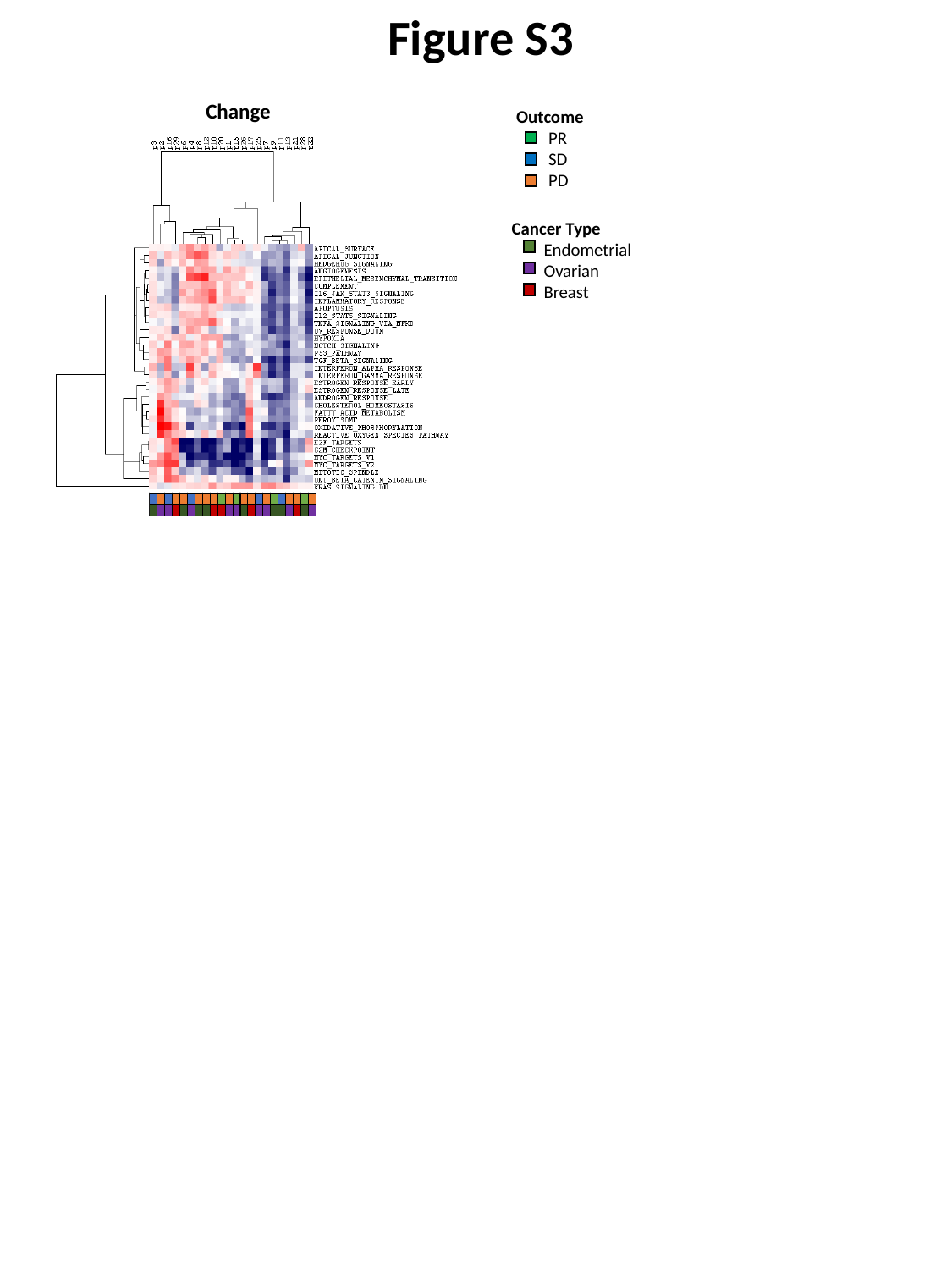

Figure S3
Change
Outcome
 PR
 SD
 PD
Cancer Type
 Endometrial
 Ovarian
 Breast

### Slide 4
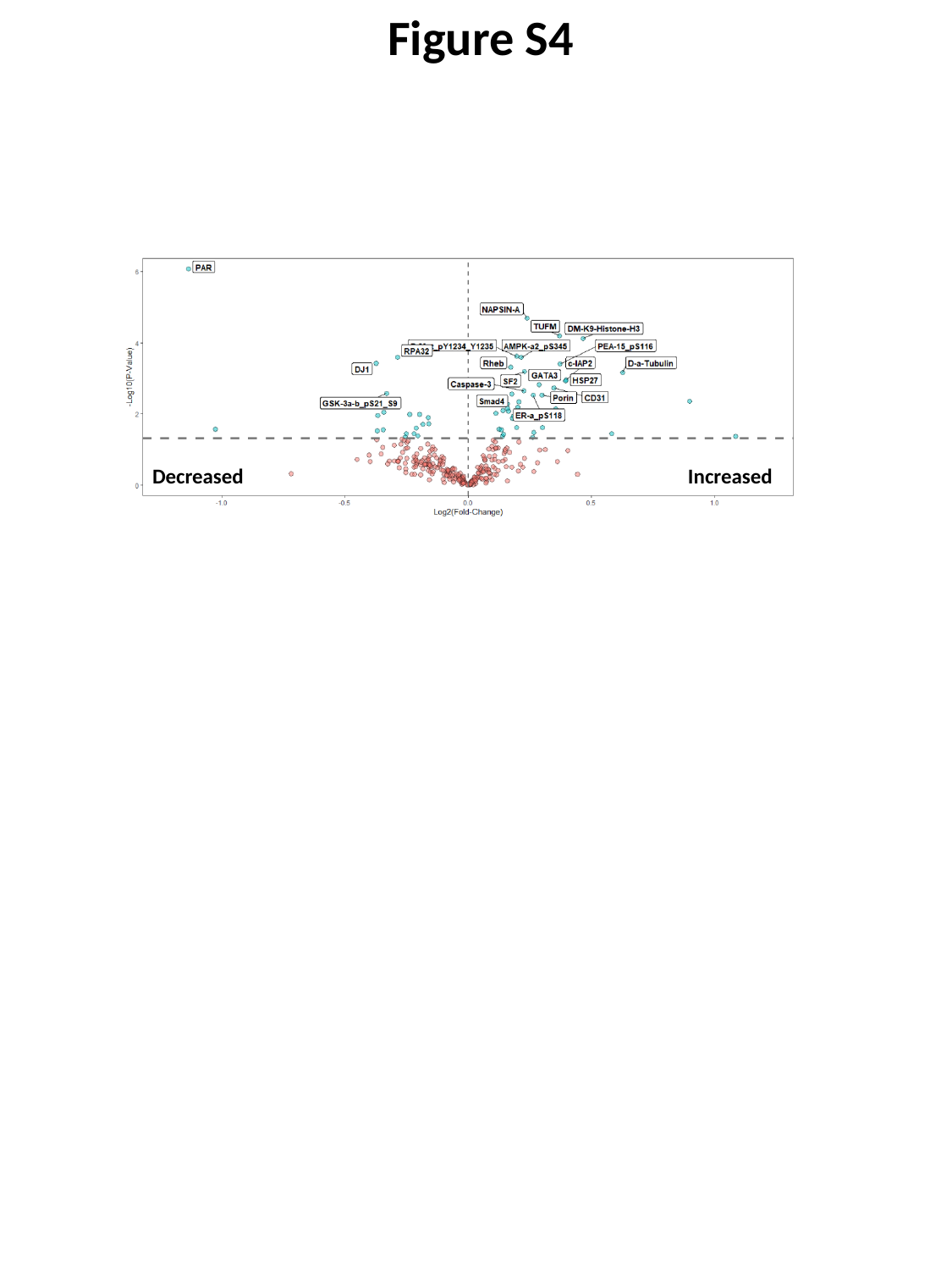

Figure S4
Decreased
Increased

### Slide 5
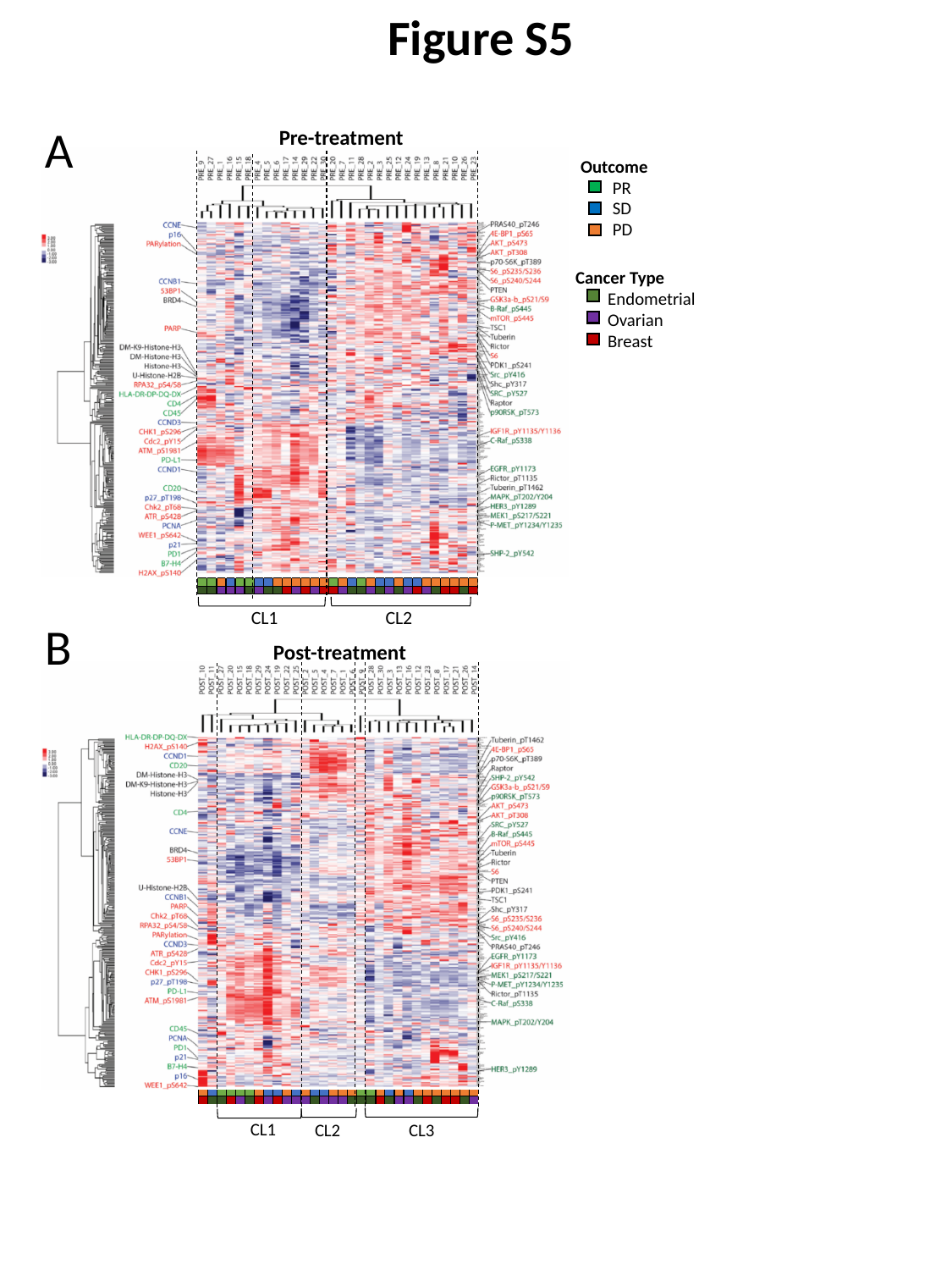

Figure S5
A
Pre-treatment
Outcome
 PR
 SD
 PD
Cancer Type
 Endometrial
 Ovarian
 Breast
CL2
CL1
B
Post-treatment
CL1
CL3
CL2

### Slide 6
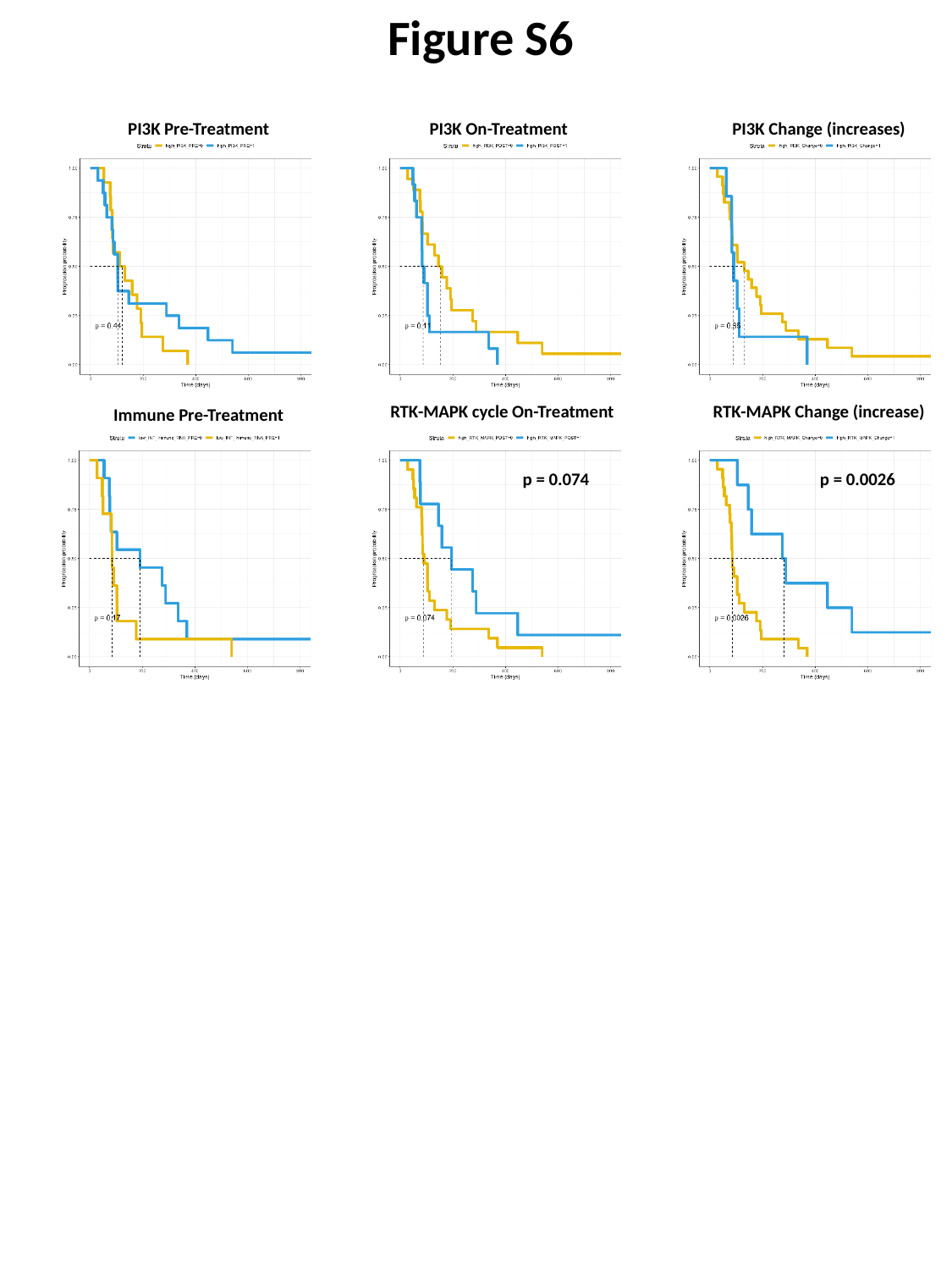

Figure S6
PI3K Pre-Treatment
PI3K On-Treatment
PI3K Change (increases)
RTK-MAPK cycle On-Treatment
RTK-MAPK Change (increase)
Immune Pre-Treatment
p = 0.074
p = 0.0026

### Slide 7
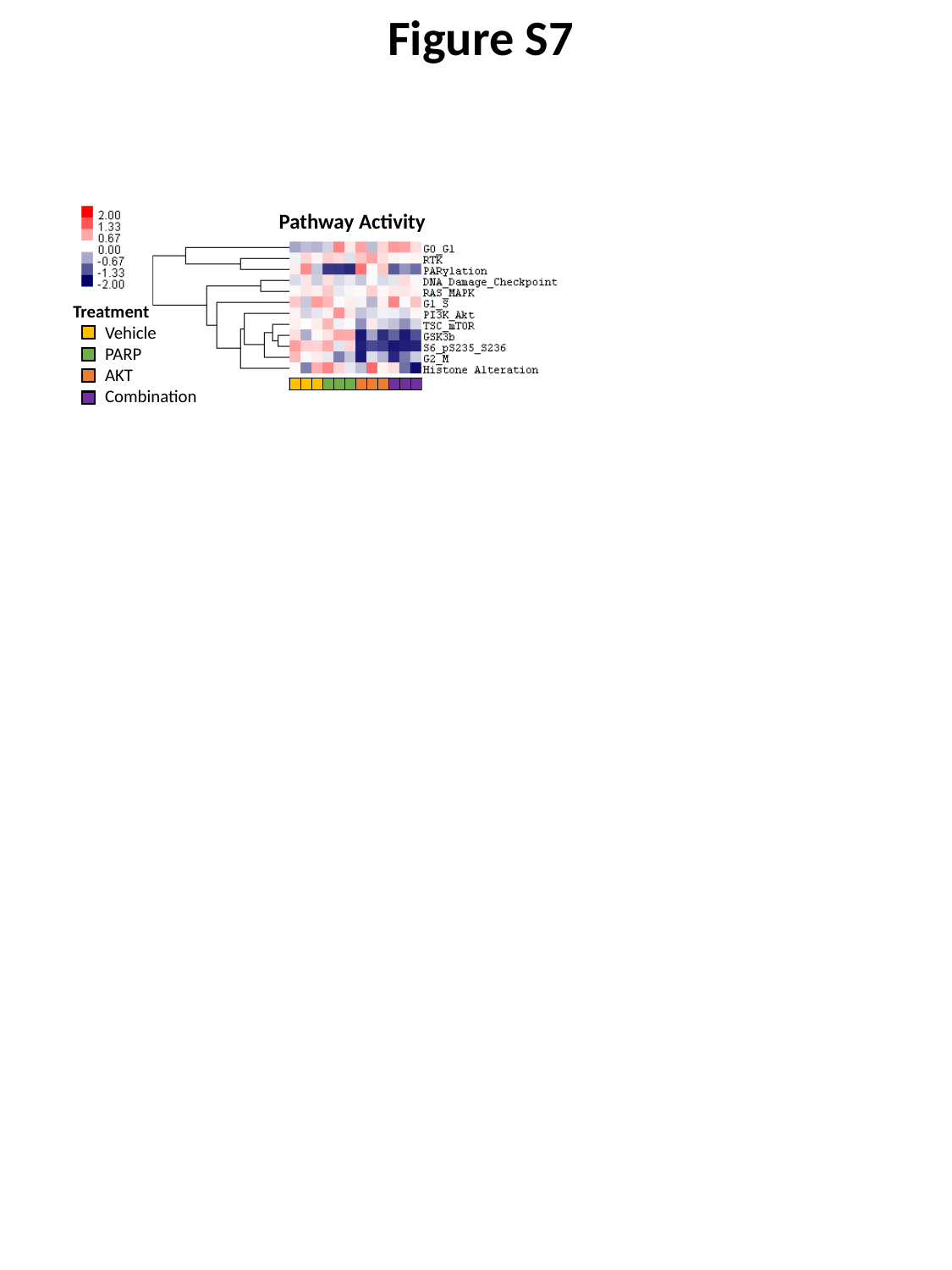

Figure S7
Pathway Activity
Treatment
 Vehicle
 PARP
 AKT
 Combination
