## Supplemental Materials for "Phase 1b dose expansion and translational analyses of olaparib in combination with the oral AKT inhibitor capivasertib in recurrent endometrial, triple negative breast, and ovarian, primary peritoneal, or fallopian tube cancer"

***Patient Population***

Eligible patients had histologically confirmed, metastatic triple negative breast cancer, recurrent endometrial adenocarcinoma (except for carcinosarcoma), recurrent high-grade serous ovarian/primary peritoneal/fallopian tube carcinoma (“ovarian cancer”), or recurrent ovarian cancer of any histology with *BRCA* mutation for whom no curative option was available. Patients with platinum-sensitive, platinum-resistant, and platinum-refractory disease were allowed. Platinum sensitivity was defined as more than six months free of relapse after last dose of platinum-based therapy. Platinum resistant and platinum-refractory disease were defined as between 3 and 6 months or less than three months free of relapse after last dose of platinum-based therapy, respectively. Measurable disease by RECIST 1.1 [1] and ECOG performance status ≤ 1 was required. Patients could have unlimited prior therapy including prior treatment with PARP, mTOR, or AKTi, as long as this therapy was not discontinued for toxicity.

Adequate bone marrow (absolute neutrophil count (ANC) ≥ 1,500/mcL, hemoglobin ≥ 10.0 g/dL, platelets ≥ 100,000/mcL), liver (total bilirubin within institutional limits, AST/ALT ≤ 2.5x institutional upper limit of normal) (ULN) in absence of liver metastases, and AST/ALT ≤ 5x institutional ULN in presence of liver metastases), and renal function (creatinine within institutional normal limits or creatinine clearance ≥ 50 mL/min/1.72 m^2^) was required. Key exclusion criteria included anti-cancer treatment or blood transfusion within 28 days of enrollment, symptomatic spinal cord compression and/or brain metastases, or residual toxicities not resolved to ≤ grade 1 (excluding alopecia). Patients with known diabetes or hyperglycemia on screening labs (HgA1C > 8% or fasting blood glucose ≥ 126mg/dL) were excluded if adequate glucose control could not be achieved with dietary measures alone. For the expansion phase, patients were required to have measurable disease accessible for biopsy. All procedures involving human participants were carried out in accordance with the Declaration of Helsinki. All subjects provided written informed consent and the study was Institutional Review Board approved.

***Assessments***

Safety assessments were performed weekly in first month and monthly thereafter. Toxicities were assessed using the Common Terminology Criteria for Adverse Events (CTCAE version 4.03). Dose limiting toxicities (DLTs) were defined as a treatment-related toxic effect occuring during the first 4 weeks of therapy. Specifically, this included 1) any non-hematologic grade 3 or 4 (excluding nausea, vomiting, dehydration, diarrhea, fatigue or asymptomatic electrolyte abnormality), 2) any grade 4, non-neutropenic, hematologic toxicity or any grade treatment- related neutropenia lasting more than 7 days, 3) any treatment-related hematologic toxicity requiring treatment delay beyond 2 weeks, 4) any grade 3 or 4 nausea, vomiting, diarrhea, dehydration or fatigue persisting greater than 72 hours despite maximally supportive care, 5) any grade 3 or 4 electrolyte abnormality (including hypocalcemia, hypokalemia, hypomagnesemia, hyponatremia, and hypophosphatemia) not corrected to ≤ grade 1 within 72 hours, and 6) any grade 3 thrombocytopenia with clinically significant bleeding.

Dose modifications and interruptions were implemented based on specific guidelines in the protocol. Broadly, adverse events that were grade 3 or more required dose interruption until resolution of toxicity to ≤ grade 1. Dose reductions were required if the adverse event did not resolve within 7 days. Delay in therapy was permitted for a maximum of 21 days. Patients with nonhematologic grade 4 toxicities were discontinued from the study. Dose re-escalation was not permitted after resolution of toxicity.

Imaging assessments were performed every 2 cycles for the first 6 months by CT or MRI of chest, abdomen, pelvis. After 6 months on treatment, response assessment was extended to every three cycles. Clinical activity was assessed by RECIST 1.1 and clinical benefit defined as objective response rate and proportion of patients progression-free at 4 months after study entry.

**Molecular analysis**

*High-depth targeted sequencing analysis*

DNA sequencing was performed in the MD Anderson Cancer Genomics Core Laboratory as previously described^21^. Genomic DNA from each frozen tissue samples was prepared and high-depth targeted sequencing (T200.2 panel) was performed. Briefly, genomic DNA were quantified by PicoGreen (Invitrogen) and quality was measured by a 2200 TapeStation system (Agilent). The sheared genomic DNA proceeded to library prep using a KAPA Hyper Prep Kit and were assessed using a TapeStation to verify DNA fragment size followed by quantification using a KAPA qPCR Quantification Kit (KAPA Biosystems). The captured libraries using biotin-labeled probe with Roche NibmleGen were sequenced using a HiSeq 2000 system (Illumina).

*Whole Exome Sequencing (WES) Mutations and Small Insertions/Deletions*

WES was obtained through standard approaches. Variants from Mutect were filtered to consider only exonic mutations, and further only nonsynonymous and stop/gain mutations^22^. Variants were filtered for a tumor allele fraction >0.05. For each sample, we assessed whether a particular gene mutation including insertions and/or deletions (in/dels) was found in the pre-treatment sample, the on-treatment sample, or both. Mutation frequencies reported in Results are tabulated by considering each gene mutation only once across pre- and on- treatment samples for each patient. Variants from Pindel are summarized by gene^23^. For *BRCA1/2* genes only, we included germline and somatic variants. We assessed whether each altered gene was associated with patient response groups. For each altered gene, we grouped mutations and in/dels (as shown in Supp Figure 1A). Fisher’s exact test of independence was used to test the relationship between gene alteration status and response. We report those genes with p-value <0.10 but note that none of the genes passes multiple testing correction when requiring a false discovery rate (FDR) <0.05.

*Comparison of Clinical Laboratory Improvement Amendments (CLIA) and WES mutations*

CLIA mutations were determined in the CLIA-accredited Molecular Diagnostics Laboratory at MDACC from patient’s archived turmor samples using routine diagnostic or therapeutic procedures. The CLIA mutation data sets were aggregated and annotated from the electronic health record at MDACC. To assess concordance of mutations called by CLIA assay and by WES, we considered both a) concordance at the gene level and b) concordance at the gene and amino acid change level. For this assessment, we checked CLIA reported mutations against the mutations called for the pre-treatment sample and the on-treatment sample. For all mutations, we evaluated functional annotation (pathogenicity) information using FASMIC^24^, OncoKB^25^, and BRCAExchange^26^. For a selected set of genes (*AKT*1/2/3, *BRCA1/2*, *HRAS*, *KRAS, MTOR, PIK3CA, PIK3R1, PTEN*, and *TP53*), we manually reviewed agreement of functional calls to provide a final functional annotation call.

*Pathway analysis*

For each patient response group (partial response: PR, stable disease: SD, and progressive disease: PD), we mapped all altered genes to Reactome^27^ pathways using the ReactomeFIViz^28^ Cytoscape^29^ application. We then assessed pathways that had an over-representation of altered genes. We required an FDR-adjusted p-value of <0.05 and more than 5 genes to be altered in each pathway. We selected only those enriched pathways that were found uniquely in one response group or in the combined PR/SD category. Because Reactome is hierarchical, we collapsed our significant pathways to parent pathways to eliminate redundancy of hit genes. Collapsed pathway categories are shown in **Supplemental Table S2** with the number of altered genes, the number of patients with an altered gene in the cohort, and the best FDR for collapsed pathways (after meeting requirement of FDR<0.05).

*RNA sequencing analysis*

RNA sequencing (RNA-seq) was performed in the MD Anderson Cancer Genomics Core Laboratory as previously described^21^. Briefly, total RNA from each frozen tissue was isolated, and the capture step was performed using biotin-labeled probes from Roche NimbleGen (Exome V3). Total RNA was assessed by 2200 TapeStation system (Agilent) and RNA was converted to double-stranded cDNA using an Ovation RNA-Seq System V2 kit (NuGEN). Sequencing proceeded as described above for the high depth sequencing analysis.

*Gene expression and pathway analysis*

Genes with less than 5 RPKM in at least 15% of the samples were filtered out. A heat map was produced by selecting all genes with a p-value ≤ 0.05 when comparing PR to PD. Hierarchical unsupervised clustering of all genes and samples was performed using publicly available Cluster 3.0 and visualized with TreeView. GSEA was performed using the reactome database and 100 permutations.

*Reverse-phase protein analysis and pathway scores*

Reverse-phase protein array was performed on protein extract as previously described^30,31^. Protein data were Z-scored, using the median and standard deviation across samples for each protein and heat maps were generated using Cluster 3.0 and TreeView. Pathway scores were calculated as previously described^10,32^. The list of predictors for each pathway is in **Supplementary Table S3**.

*PDX model*

TNBC PARP-resistant PDX model (BCX.099) was implanted into NSG immunodeficient mice. Mice were treated with vehicle, olaparib (50mg/kg QC continuous), capivasertib (100 mg/kg bid, 4 days on, 3 days off) or their combination (same dose, co-formulation) for five cycles. Tumors were measured once a week and were harvested after the fifth cycle for RPPA analysis. All animal experiments were performed in accordance with Institutional Animal Care and Use Committee (IACUC) protocols.
