## Supplemental tables for "Phase 1b dose expansion and translational analyses of olaparib in combination with the oral AKT inhibitor capivasertib in recurrent endometrial, triple negative breast, and ovarian, primary peritoneal, or fallopian tube cancer"

**Supplemental table S1: Schedule of patient reported outcomes instruments**

| **Instrument** | **PRO Type** | **Assessment time points^5^** |
| --- | --- | --- |
| MDASI-Ovarian Cancer^1^ | Symptom burden | Baseline, every week for 2 cycles, then every cycle, and at time patient comes off study |
| FACT-EGFR-18^2^ | Skin and dermatologic symptoms | Baseline, every 2 cycles, and at time patient comes off study |
| FACT-Ovary^3^ | Quality of life | Baseline, every 2 cycles, and at time patient comes off study |
| Additional symptoms and symptom burden change | Symptoms that may not have been captured on existing surveys, and symptom burden change over the past cycle | Every 2 cycles |
| PHQ-9^4^ and GAD-7^5^ | Anxiety and depression | Baseline, every 2 cycles, and at time patient comes off study |
| FACIT-TS-PS^6^ | Satisfaction with care | Baseline and and at time patient comes off study |
| EQ-5D-5L^7,8^ | Health outcome, health perception | Baseline, every 2 cycles, and at time patient comes off study |

**Supplemental Table S2**: **Enriched pathways by response group.** Pathways have been collapsed to parent pathways to remove redundancy. FDR shown in table is minimum FDR of collapsed pathways.

| **Pathway** | **Group** | **Genes Altered in Pathway** | **FDR*** | **Patients** | **N Genes** | **N Patients** |
| --- | --- | --- | --- | --- | --- | --- |
| Cell Cycle Checkpoints | PR | CDC20, TOP3A, TP53, AURKB, TP53BP1, CDKN2A, MDM2, ATM | 0.0010576 | p18;p28;p15;p20;p27;p9 | 8 | 6 |
| Signaling by FGFR | PR | FGF5, PPP2R1A, HRAS, PIK3CA, SRC, PIK3R1, SRC, FGFR3 | 2.59E-06 | p18;p27;p28;p9;p20 | 7 | 5 |
| Signaling by NTRKs | PR | HRAS, PIK3CA, SRC, PIK3R1, NTRK2, NTRK3 | 2.58E-04 | p18;p20;p9;p28 | 6 | 4 |
| Cell Cycle, Mitotic | PR, SD | CDC20, PPP2R1A, AKT1, POLE, TP53, AURKB, CDKN2C, CDKN2A, RB1, TOP2A, CDKN1A, CDC27, EP300, STAG2 | 0.0022923 | p18;p3;p9;p19;p15;p16;p20;p24;p25;p27;p28;p4;p5;p11 | 14 | 14 |
| Signaling by Nuclear Receptors | SD | FOXA1, EP300, RUNX1, RARA, CREBBP, STAG2 | 0.002256 | p16;p25;p4;p3;p5;p11;p24 | 6 | 7 |
| Axon guidance | PD | RET, GSK3B, EPHA5, PIK3CA, MET, EPHA2, PIK3R1, EGFR, KRAS | 0.0295 | p12;p26;p2;p17;p21;p29;p6;p23;p14;p22;p1 | 9 | 11 |
| Cytokine Signaling in Immune System | PD | AKT1, TP53, PIK3R1, GATA3, STAT3, VEGFA, SYK, PIK3CA, CSF1R, SMARCA4 | 0.0016214 | p14;p21;p23;p29;p8;p10;p12;p13;p2;p22;p26;p30;p6;p7p14;p21;p23;p29;p8;p17;p26;p6;p10;p12;p13;p2;p22;p30;p7;p1 | 10 | 17 |
| MAPK family signaling cascades | PD | RET, FGF5, PDGFRA, KIT, MET, EGFR, NF1, KRAS, FGFR3, FGFR2 | 8.75E-05 | p12;p26;p8;p13;p30;p29;p22;p10;p1;p6;p14 | 10 | 11 |
| Signaling by ERBB4 | PD | NCOR1, PIK3CA, PIK3R1, EGFR, ESR1, KRAS | 4.79E-05 | p22;p17;p21;p26;p29;p6;p14;p23;p1;p12 | 6 | 10 |
| Signaling by NOTCH | PD | EP300, TP53, NOTCH2, NOTCH3, NOTCH1, CREBBP, FLT4, AKT1, | 1.59E-05 | p26;p30;p6;p7;p10;p12;p13;p2;p22;p23;p29;p8;p1;p14;p21p26;p14;p21;p23;p29;p8;p30;p6;p7;p12;p1;p22 | 8 | 16 |
| Signaling by WNT | PD | GSK3B, AKT1, EP300, KMT2D, TERT, CREBBP, SMARCA4, APC | 4.17E-04 | p26;p14;p21;p23;p29;p8;p30;p6;p7;p12;p2;p22;p1 | 8 | 13 |
| Transcriptional regulation by the AP-2 (TFAP2) family of transcription factors | PD | EP300, KIT, EGFR, CREBBP, ESR1, VEGFA | 1.33E-05 | p26;p30;p6;p7;p13;p22;p1;p14;p21;p23;p8;p17;p2 | 6 | 13 |
| Transcriptional regulation by TP53 | PD | BRCA1, MSH2, FANCD2, TP53, ERCC3, CDK12 | 1.31E-04 | p10;p30;p7;p6;p12;p13;p2;p22;p23;p26;p29;p8;p21 | 6 | 13 |

**Supplemental Table 3. Pathway score predictors used with the patient samples**

| **Pathway** | **Predictor** | **Weight** | **Count** |
| --- | --- | --- | --- |
| DNA_Damage_Checkpoint | ATM_pS1981 | 1 | 1 |
| DNA_Damage_Checkpoint | ATR_pS428 | 1 | 1 |
| DNA_Damage_Checkpoint | cdc2_pY15 | 1 | 1 |
| DNA_Damage_Checkpoint | Chk1_pS296 | 1 | 1 |
| DNA_Damage_Checkpoint | Chk2_pT68 | 1 | 1 |
| DNA_Damage_Checkpoint | Wee1_pS642 | 1 | 1 |
| G0_G1 | BRD4 | -1 | 1 |
| G0_G1 | Cyclin-B1 | -1 | 1 |
| G0_G1 | 14-3-3-beta | 1 | 1 |
| G0_G1 | 53BP1 | 1 | 1 |
| G0_G1 | Cyclin-D1 | 1 | 1 |
| G0_G1 | p27_pT198 | 1 | 1 |
| G0_G1 | p21 | 1 | 1 |
| G1_S | ATM_pS1981 | -1 | 1 |
| G1_S | CD134 | -1 | 1 |
| G1_S | GATA6 | -1 | 1 |
| G1_S | 53BP1 | 1 | 1 |
| G1_S | BRD4 | 1 | 1 |
| G1_S | Cyclin-E1 | 1 | 1 |
| G1_S | PKM2 | 1 | 1 |
| G2_M | Cyclin-B1 | 1 | 1 |
| G2_M | PLK1 | 1 | 1 |
| G2_M | CDK1 | 1 | 1 |
| G2_M | cdc25C | 1 | 1 |
| G2_M | Rb_pS807_S811 | 1 | 1 |
| GSK3b | GSK-3a-b_pS21_S9 | 1 | 1 |
| Histone_Alteration | Histone-H3 | 1 | 1 |
| Histone_Alteration | U-Histone-H2B | 1 | 1 |
| Histone_Alteration | DM-Histone-H3 | 1 | 1 |
| Histone_Alteration | DM-K9-Histone-H3 | 1 | 1 |
| Histone_Alteration | H2AX_pS140 | 1 | 1 |
| Immune | CD4 | 1 | 1 |
| Immune | CD45 | 1 | 1 |
| Immune | Lck | 1 | 1 |
| Immune | ZAP-70 | 1 | 1 |
| Immune_Checkpoint | B7-H4 | 1 | 1 |
| Immune_Checkpoint | CD134 | 1 | 1 |
| Immune_Checkpoint | PD-1 | 1 | 1 |
| Immune_Checkpoint | PD-L1 | 1 | 1 |
| PARylation | PAR | 1 | 1 |
| PI3K_Akt | INPP4b | -1 | 1 |
| PI3K_Akt | PTEN | -1 | 1 |
| PI3K_Akt | Akt_pS473 | 0.5 | 0.5 |
| PI3K_Akt | Akt_pT308 | 0.5 | 0.5 |
| PI3K_Akt | GSK-3a-b_pS21_S9 | 1 | 1 |
| PI3K_Akt | p27_pT198 | 1 | 1 |
| PI3K_Akt | PRAS40_pT246 | 1 | 1 |
| PI3K_Akt | Tuberin_pT1462 | 1 | 1 |
| RAS_MAPK | B-Raf_pS445 | 1 | 1 |
| RAS_MAPK | c-Jun_pS73 | 1 | 1 |
| RAS_MAPK | C-Raf_pS338 | 1 | 1 |
| RAS_MAPK | JNK_pT183_Y185 | 1 | 1 |
| RAS_MAPK | MAPK_pT202-Y204 | 1 | 1 |
| RAS_MAPK | MEK1_p_S217-S221 | 1 | 1 |
| RAS_MAPK | p38_pT180_Y182 | 1 | 1 |
| RAS_MAPK | p38-MAPK | 1 | 1 |
| RAS_MAPK | p90RSK_pT573 | 1 | 1 |
| RAS_MAPK | YB1_pS102 | 1 | 1 |
| RTK | Src_pY416 | 0.5 | 0.5 |
| RTK | Src_pY527 | 0.5 | 0.5 |
| RTK | Axl | 1 | 1 |
| RTK | c-Met_pY1234_Y1235 | 1 | 1 |
| RTK | EGFR_pY1173 | 1 | 1 |
| RTK | HER2_pY1248 | 1 | 1 |
| RTK | HER3_pY1289 | 1 | 1 |
| RTK | IGF1R_pY1135_Y1136 | 1 | 1 |
| RTK | IRS1 | 1 | 1 |
| RTK | Shc_pY317 | 1 | 1 |
| RTK | SHP-2_pY542 | 1 | 1 |
| S6 | S6 | 1 | 1 |
| TSC_mTOR | S6_pS235_S236 | 0.5 | 0.5 |
| TSC_mTOR | S6_pS240_S244 | 0.5 | 0.5 |
| TSC_mTOR | 4E-BP1_pS65 | 1 | 1 |
| TSC_mTOR | mTOR_pS2448 | 1 | 1 |
| TSC_mTOR | p70-S6K_pT389 | 1 | 1 |
| TSC_mTOR | Rb_pS807_S811 | 1 | 1 |
| TSC_mTOR | Rictor_pT1135 | 1 | 1 |

**Supplemental Table 3. Pathway score predictors used with the PDX model**

| **Pathway** | **Predictor** | **Weight** | **Count** |
| --- | --- | --- | --- |
| DNA_Damage_Checkpoint | ATM_pS1981 | 1 | 1 |
| DNA_Damage_Checkpoint | ATR_pS428 | 1 | 1 |
| DNA_Damage_Checkpoint | cdc2_pY15 | 1 | 1 |
| DNA_Damage_Checkpoint | Chk1_pS296 | 1 | 1 |
| DNA_Damage_Checkpoint | Chk2_pT68 | 1 | 1 |
| DNA_Damage_Checkpoint | Wee1_pS642 | 1 | 1 |
| G0_G1 | BRD4 | -1 | 1 |
| G0_G1 | Cyclin-B1 | -1 | 1 |
| G0_G1 | 14-3-3-beta | 1 | 1 |
| G0_G1 | 53BP1 | 1 | 1 |
| G0_G1 | Cyclin-D1 | 1 | 1 |
| G0_G1 | p27_pT198 | 1 | 1 |
| G0_G1 | p21 | 1 | 1 |
| G1_S | ATM_pS1981 | -1 | 1 |
| G1_S | CD134 | -1 | 1 |
| G1_S | 53BP1 | 1 | 1 |
| G1_S | BRD4 | 1 | 1 |
| G1_S | PKM2 | 1 | 1 |
| G2_M | Cyclin-B1 | 1 | 1 |
| G2_M | PLK1 | 1 | 1 |
| G2_M | CDK1 | 1 | 1 |
| G2_M | cdc25C | 1 | 1 |
| G2_M | Rb_pS807_S811 | 1 | 1 |
| GSK3b | GSK-3a-b_pS21_S9 | 1 | 1 |
| Histone_Alteration | Histone-H3 | 1 | 1 |
| Histone_Alteration | DM-Histone-H3 | 1 | 1 |
| Histone_Alteration | DM-K9-Histone-H3 | 1 | 1 |
| PARylation | PAR | 1 | 1 |
| PI3K_Akt | INPP4b | -1 | 1 |
| PI3K_Akt | PTEN | -1 | 1 |
| PI3K_Akt | Akt_pS473 | 0.5 | 0.5 |
| PI3K_Akt | Akt_pT308 | 0.5 | 0.5 |
| PI3K_Akt | GSK-3a-b_pS21_S9 | 1 | 1 |
| PI3K_Akt | p27_pT198 | 1 | 1 |
| PI3K_Akt | PRAS40_pT246 | 1 | 1 |
| PI3K_Akt | Tuberin_pT1462 | 1 | 1 |
| RAS_MAPK | B-Raf_pS445 | 1 | 1 |
| RAS_MAPK | c-Jun_pS73 | 1 | 1 |
| RAS_MAPK | C-Raf_pS338 | 1 | 1 |
| RAS_MAPK | JNK_pT183_Y185 | 1 | 1 |
| RAS_MAPK | MAPK_pT202-Y204 | 1 | 1 |
| RAS_MAPK | MEK1_p_S217-S221 | 1 | 1 |
| RAS_MAPK | p38_pT180_Y182 | 1 | 1 |
| RAS_MAPK | p38-MAPK | 1 | 1 |
| RAS_MAPK | p90RSK_pT573 | 1 | 1 |
| RAS_MAPK | YB1_pS102 | 1 | 1 |
| RTK | Src_pY416 | 0.5 | 0.5 |
| RTK | Src_pY527 | 0.5 | 0.5 |
| RTK | Axl | 1 | 1 |
| RTK | c-Met_pY1234_Y1235 | 1 | 1 |
| RTK | EGFR_pY1173 | 1 | 1 |
| RTK | HER2_pY1248 | 1 | 1 |
| RTK | HER3_pY1289 | 1 | 1 |
| RTK | IGF1R_pY1135_Y1136 | 1 | 1 |
| RTK | IRS1 | 1 | 1 |
| RTK | Shc_pY317 | 1 | 1 |
| RTK | SHP-2_pY542 | 1 | 1 |
| S6_pS235_S236 | S6_pS235_S236 | 1 | 1 |
| TSC_mTOR | S6_pS235_S236 | 0.5 | 0.5 |
| TSC_mTOR | S6_pS240_S244 | 0.5 | 0.5 |
| TSC_mTOR | 4E-BP1_pS65 | 1 | 1 |
| TSC_mTOR | mTOR_pS2448 | 1 | 1 |
| TSC_mTOR | p70-S6K_pT389 | 1 | 1 |
| TSC_mTOR | Rb_pS807_S811 | 1 | 1 |
| TSC_mTOR | Rictor_pT1135 | 1 | 1 |
